# Phylogenetic Insights into SARS-CoV-2 Introductions and Spread in Georgia

**DOI:** 10.64898/2026.03.23.26349139

**Authors:** G. Veytsel, L. Lyu, G. Stott, L. Carmola, H. Dishman, J. Bahl

## Abstract

The spread of successive novel COVID-19 variants presented a challenge for outbreak surveillance, epidemiology, and emergency responses. Monitoring the emergence and spread of SARS-CoV-2 variants is essential to allocate limited public health resources and optimize control efforts. Global collaboration among the scientific community enabled large-scale viral surveillance and sequencing efforts. However, translating these vast datasets into actionable public health inferences requires rapid statistical methodologies, scalable workflows, and robust frameworks.

In this study, we focused on the Delta epidemic wave in Georgia by applying a hybrid maximum likelihood (ML) and Bayesian phylodynamic approach. We characterized the Delta variant introduction to Georgia and its subsequent local spread. Our analysis of 9,783 Delta sequences collected between August 1, 2020 and January 25, 2022 detected at least 344 introductions into Georgia, resulting in 34 highly-supported local clusters. On average, clusters circulated for one month before the earliest detected sequence, highlighting critical delays in detection. While most clusters remained small, a few introduction events led to large, sustained outbreaks. We jointly inferred the statewide transmission network, estimated from all locally circulating clusters with a modified Bayesian discrete trait phylogeographic reconstruction of statewide health districts. We showed that South Central, Georgia was a major source of transmission, despite having smaller numbers of infected people, compared to major metropolitan areas.

Our study addresses the urgent need for methodologies and data-driven recommendations for public health practice, particularly given large, dynamic, and integrated datasets. By identifying key geographic sources and sinks of transmission, our findings can guide resource allocation and prepare for future epidemics among high-risk populations. Additionally, by characterizing introduction events, local circulation, and detection lags, we highlight critical gaps in surveillance. These gaps can inform outbreak investigation and response, such as targeted contact tracing and testing.

## Introduction

SARS-CoV-2, the causative agent of COVID-19, is responsible for a global pandemic that caused significant morbidity and mortality worldwide along with substantial societal and economic costs (1,2). The first known COVID-19 case in the state of Georgia was confirmed on March 2, 2020, in Fulton County (3). Since, epidemic waves of COVID-19 have been associated with the emergence of novel variants of concern (VOCs) (4) that have increased the risk to public health (e.g., increased virulence, transmissibility, and immune evasion), such as Alpha, Delta, and Omicron. Consequently, it has become essential to monitor the emergence and spread of SARS-CoV-2 variants. Global collaboration across the scientific community has resulted in immense efforts in viral surveillance and sequencing to better understand COVID-19 epidemiology, including detection of clusters of transmission in various settings, investigation of outbreaks, and identification of introductions, key sources/sinks, and superspreading events (5). To date, there are over 15 million SARS-CoV-2 sequences available in the Global Initiative on Sharing All Influenza Data (GISAID) (6). However, there remains a need for rapid statistical methodologies, scalable workflows, and robust frameworks to translate these data into actionable public health inferences.

Substantial bioinformatics tools have been developed or updated to store, assess, and analyze massive SARS-CoV-2 data in nearly all aspects of SARS-CoV-2 research (7). While this wealth of data provides an unprecedented opportunity to study epidemic transmission dynamics, these large and dynamically growing datasets have exposed the computational limitations of existing molecular epidemiological approaches (8). Bayesian phylodynamics, the gold standard approach to infer the dispersal of viral lineages, cannot tackle large datasets in a reasonable time frame.

The computational power, memory requirements, and technical expertise may also not be readily available in resource-limited settings. To address the practical limitations of Bayesian phylodynamics, alternative pipelines have been proposed to reduce computation time by combining non-clocklike (fast) maximum-likelihood (ML) and Bayesian phylodynamic estimation (9).

There have been a few epidemiological studies that focus on the early months of the COVID-19 pandemic in Georgia. A phylogenetic study of 108 Georgia sequences obtained through the end of March 2020 identified 19 introduction events into the state, revealing an important contribution of external introduction events in establishing local transmission chains in Georgia (10). An early modeling study of case surveillance data found that superspreading was an important and pervasive feature throughout the COVID-19 pandemic in Georgia and may play a critical role in driving large outbreaks in rural areas (11). Even within the same geographic region, we expect that invasion dynamics and local spread may be substantially different between the early epidemic and ensuing epidemic waves due to changes in population immunity and vaccination coverage over time, fluctuations in interventions and restrictions, and differences in characteristics between VOCs.

The Delta variant (B.1.617.2 and AY* sub-lineages) was first recognized in India in early 2021 and displaced Alpha as the dominant variant in mid-2021, leading to a resurgence of COVID-19 cases in many countries (12). Delta is associated with increased transmissibility and greater disease severity due to immune evasion and higher virus loads resulting from the antagonism of innate immunity (12,13). Little is known about how Delta invaded Georgia and subsequent local spread. SARS-CoV-2 can exhibit substantial transmission heterogeneity. Superspreader events have revealed that a single introduction event can lead to sustained community transmission with little broader impact or be exported with extensive regional, national, and international spread (10,11,14,15).

In this study, we characterize the invasion of Delta into Georgia. To focus on lineages that contributed most to the epidemic in Georgia, we track lineages that led to local transmission and genetic diversity in the state. Specifically, Delta lineages that had been introduced into Georgia and seeded an outbreak with onward local spread. These represent local clusters in the state, i.e., genetically similar infections that are related through a series of recent transmission events.

Clusters can be used to characterize the structure of an epidemic driven by repeated introductions (16). Thus, we use local clusters to reveal the complex structure of Delta transmission in Georgia. We build on previous methods to (1) detect introduction events of Delta into Georgia and identify clusters that arose through independent introductions using an ML approach and (2) discover the statewide Delta transmission network in Georgia using Bayesian phylogeographic reconstruction across the local clusters. We characterize the number and timing of introductions, describe the onward spread (do introductions generally result in limited spread or sustained transmission?), infer source-sink dynamics within Georgia clusters, and estimate the overall transmission network between public health districts. In conclusion, we provide a rapid and robust statistical framework and actionable inferences that can be used for public health investigation, response, and policy during future outbreaks.

## Materials and Methods

### Data

Whole genome sequences of SARS-CoV-2 obtained from human COVID-19 cases in Georgia were downloaded from the Global Initiative on Sharing Avian Influenza Data-EpiCoV (GISAID-EpiCoV) platform (https://www.gisaid.org/) (6). Only Delta (B.1.617.2 and AY) sequences submitted up to May 10, 2023 with complete genomes (greater than 29,000 nucleotides) and complete collection dates were included. To ensure high quality sequences, a high coverage filter was applied, which GISAID defines as entries with less than 1% of undefined bases (Ns) and no insertion/deletions unless verified by submitter. Genomes were linked to location metadata provided by the Georgia Department of Public Health. The zipcodeR package (18) was used to assign zip codes to county. All county designations were transformed into public health districts, an administrative boundary for health policy and funding, based on the following designations: https://dph.georgia.gov/public-health-districts. Sequences with an unidentified public health district were excluded from the final dataset (n = 20,674). To inform the subsampling strategy, all PCR confirmed and antigen-positive SARS-CoV-2 case data, by county and week, were downloaded from the Georgia Department of Health (https://dph.georgia.gov/covid-19-status-report). Sequences were subsampled proportionally to the number of cases in each public health district in each week, with a maximum of 5,000 sequences. This strategy aims to prioritize sequences that reflect the epidemic size over time and space, while mitigating potential sequencing biases. To generate a summary table of key demographic characteristics of these public health districts, data was compiled from the U.S. Census (19) and National Institute on Minority Health and Health Disparities (20).

To build a worldwide contextual dataset, a random sample of 50,000 genomes was downloaded from all global sequences available in GISAID as of September 15, 2024. Only Delta sequences submitted up to December 11, 2021, the last week of the Delta wave in Georgia, were queried. Previous criteria were applied: human host, high coverage, and complete collection date (n=3,160,077). Nextstrain’s ncov pipeline v13 (github.com/nextstrain/ncov) was used to filter and align/mask genomic sequences (21). Contextual sequences were grouped by collection year, week, and region in order to generate a spatially and temporally representative subsample. To keep a 1:1 ratio of focal to contextual sequences, a maximum of 5,000 sequences were selected to be as close as possible to the focal Georgia sequences using priority = “proximity”. A crowding penalty of 0.1 was applied when choosing contextual samples for a focal set to reduce the number of genetically identical strains that are chosen and allows for more diversity represented on the tree. Genomes were aligned against the Wuhan reference sequence (GenBank accession MN908947). The final alignment was masked at the beginning (first 100 bases) and end (last 50 sites), regions of the genome that are difficult to sequence accurately. Positions 21987 (problematic ARTIC V3 primer site) and 21846 (often artifactually reverted in Delta) were also masked to improve the quality of the alignment.

*Initial maximum likelihood phylogenetic analysis to identify Georgia clusters*. Following previous phylogenetic studies of SARS-CoV-2 (14,17,22–24), ancestral state reconstruction was used to identify introduction events into Georgia and resulting Georgia transmission clusters. An evolutionary tree was constructed under a maximum likelihood framework and branch support values were estimated using IQ-Tree v2.2 (25). The best-fit model for tree construction was determined to be a Generalized Time Reversible (26) (GTR) model with empirical base frequencies (+F), allowing for a proportion of invariable sites (+I), and discrete Gamma model with default 4 rate categories (+G4) (27) via ModelFinder (28). Branch support was assessed using ultrafast bootstrap (UFBoot2) approximation (29,30) (1000 replicates) and single branch support tests, like the SH-like approximate likelihood ratio test (SH-LRT) (31) (1000 replicates) and the approximate Bayes test (aBayes) (32). The estimated divergence tree was input into TreeTime (33) for the estimation of rates and dates by simple least-squares regression. Outlier sequences that do not follow the molecular clock model can skew the estimation of substitution rates. From the initial tree, tips that deviated by > 8 interquartile ranges in a root-to-tip by time regression were removed. To time scale the phylogenetic tree, Wuhan/Hu-1/2019 (https://www.ncbi.nlm.nih.gov/nuccore/MN908947) was used as the root and the clock rate was fixed to 8 x 10^-4^ substitutions/site/year, per the early estimated rate of evolution (34). A table containing full posterior distributions for the inferred dates of internal nodes was also generated. TreeTime’s mugration model was applied to reconstruct the relative probability of a binary discrete character state (Georgia or non-Georgia) at every internal node of the time-scaled tree. SARS-CoV-2 sequences are highly similar, often resulting in short branch lengths, indicating lack of genetic divergence, and polytomies, indicating unresolved relationships; in cases of zero-length branches, we added a small constant (1E-8) in R (R Core Team 2021). We estimated the cumulative number of introductions (transitions from a non-Georgia node to a Georgia node/tip) based on the ancestral state reconstruction of internal nodes. To approximate how long Delta was spreading in the state prior to detection in sequencing data, we quantified the gap between the estimated time to most recent common ancestor (TMRCA) for each introduction event and the earliest sampled Georgia sequence. We defined highly supported Georgia clusters as monophyletic clades that descended from independent introduction events in Georgia with sufficient branch support (SH-aLRT ≥ 80%, aBayes ≥ 95%, and UFBoot ≥ 95%) (25) and at least 3 Georgia sequences. The resulting 34 clusters were used to characterize introduction events. The date of the non-Georgia node was used as the TMRCA of the introduction.

*Bayesian phylogenetic analysis to produce an empirical set of trees for each Georgia cluster*. The clock signal of the maximum likelihood tree was investigated with Tempest v1.5.3 (35) (Supplementary Figure 1). For subsequent discrete trait analyses, we generated an empirical set of trees from the posterior distribution of each cluster in BEAST v1.10.4 (36). To increase statistical power to infer rates, we excluded clusters that were comprised of fewer than five sequences or was made up of just one public health district, resulting in 24 clusters. The selected nucleotide substitution model (GTR+F+I+G4) and clock rate prior (8 x 10^-4^ substitutions/site/year) were consistent with input from the preliminary maximum likelihood analysis. Additionally, an uncorrelated relaxed lognormal clock model allowed for evolutionary rate variation across branches and an exponential growth coalescent tree prior modelled population size changes through time. Due to the small sample sizes in some clusters, a mean root height was specified (mean 1.0 years) with a wide standard deviation of 1.0 to support convergence. Three independent Markov Chain Monte Carlo (MCMC) runs of 100 million chain length were performed, sampling every 10,000 states. Convergence and mixing were verified using Tracer v1.7 (37) and tree files were combined in LogCombiner (36) with a burn-in of 10%. *Individual discrete trait analyses to infer Markov jumps and rewards*. Discrete trait analyses for each cluster were conducted using a continuous-time Markov chain (CTMC) model to infer transitions (Markov jumps) between public health districts along phylogenetic branches and the waiting time (Markov rewards) spent in each public health district between transitions. Empirical sets of trees were resampled to 1,000 trees for each cluster to accommodate for the memory requirements to infer complete jump history. Three independent MCMC runs of 100 million chain length were performed for each cluster, sampling every 10,000 states. Following examination in Tracer (37), log and tree files were each combined in LogCombiner (36) with a burn-in of 10%. Markov jumps were extracted from the complete jump history using a Perl script (38) to retrieve the date of each jump for each pair of public health districts from our complete jump history file and visualized as heatmaps built using ggplot2 in R (39,40).

Although a public health district may have the highest number of transitions to a particular sink, this alone does not confirm its role as a primary source. Beyond absolute counts, it is important to assess whether transitions to a given sink occur more frequently than expected relative to other possible sinks. This approach accounts for overall migration patterns and prevents misinterpretation of high transition volume as disproportionate influence. To further refine these source-sink inferences, we also compared potential sources for each sink, determining whether one district contributed more frequently than others. We quantified this using the relative risk of a public health district as a viral source for a particular sink (compared to other sinks) and assessed its potential for enhanced risk relative to other sources.

For clusters that included all the public health districts, we estimated the average number of transitions over time and quantified the relative risk of each district as a viral source, following the approach described in Bahl et al. (2016) (41). For each combination of public health districts, we constructed contingency tables summarizing the total number of transitions into and out of each district at each Bayesian MCMC step. Relative risk was calculated as the ratio of the proportion of the times a district serves as a source for the given sink versus the proportion of times it serves as a source for all other sinks. These ratios were calculated for each pair of public health districts per MCMC step and averaged across all steps to incorporate phylogenetic uncertainty. Relative risks (RR) with a Highest Density Interval (HDI) excluding 1 were considered significant. For each sink, heterogeneity among potential sources was examined: non-overlapping HDI intervals across sources indicated significantly different contributions to the sink (i.e., enhanced risk).

*Bayesian joint estimation across Georgia clusters infer rates*. Empirical sets of trees generated for each cluster in the previous step were resampled to 540 trees to accommodate for the high memory requirements in inferring rates across many empirical sets of trees. Following the approach described in Fujimoto et al., we jointly estimated a single phylodynamic discrete trait model to all clusters (42). Briefly, rather than one matrix for a tree containing all clusters, a single rate matrix was applied independently across all clusters. This allowed the statewide transmission network for Delta to be inferred, which demonstrated how connected public health districts are to each other, as well as how an outbreak in Georgia is connected to the pandemic out of state. Since the number of taxa and distribution of public health districts vary among individual clusters, a model averaging approach was applied to assess the overall epidemic spread in Georgia. This joint estimation approach assumes that the underlying mechanism of spatial diffusion remains consistent across clusters.

The social network was inferred using Bayesian Stochastic Search Variable Selection (BSSVS), which identified parameters with significantly non-zero transition rates in order to capture location exchange rates that adequately explain the diffusion process (43). By specifying an asymmetric CTMC model, discrete trait analysis enabled estimation of migration rates of viral lineages between geographic locations and reconstruction of the geographic locations of ancestral lineages. This is essential in identifying key sources and sinks of transmission. Three independent MCMC runs of 100 million chain length were performed, sampling every 10,000 states. Following examination in Tracer (37), log and tree files were each combined in LogCombiner with a burn-in of 10%. Bayes Factor (BF) values were calculated using Spread3 (Spatial Phylogenetics Reconstruction of Evolutionary Dynamics using Data-Driven Documents (D3)) (44) to analyze the support for each pairwise rate of diffusion between locations. To assess the level of support, we used common thresholds in the field: No support BF < 3; substantial support: BF = 3-10; strong support: BF = 11-30; very strong support: BF = 31-100; decisive support: BF > 100 (45,46). A maximum clade credibility (MCC) tree was obtained using TreeAnnotator v.1.10.4 (36) for each cluster and visualized in R.

To examine our assumption about the consistent underlying spatial diffusion across clusters, we also examined concordance between jointly inferred rates across all clusters and the rates of each cluster estimated independently. Is there overall agreement between the joint analysis and individual clusters?

Lastly, because this approach is dependent on the initial time-scaled ML tree, we also assessed the robustness of our results to resampling. Changing the initial starting seed upon subsampling, we generated four additional focal datasets. We reran our entire pipeline from subsampling of contextual sequences in Nextstrain to joint estimation. Across these five replicates, we compared the inferred number of introductions, the number of clusters identified, the size of the clusters, and the inferred statewide transmission network. We reported transmission rates identified by all subsamples to ensure that our inferences are robust and not sensitive to the resampling.

## Results

### SARS-CoV-2 cases across Public Health Districts in Georgia

The first Delta sequence was collected in Georgia on April 9, 2021 and the last on March 10, 2022. The Delta variant was the dominant circulating SARS-CoV-2 variant for 24 weeks (June 27, 2021 – December 11, 2021) (Supplementary Figure 2). Georgia is divided into 18 public health districts (Figure 1, Supplementary Figure 3); each encompasses one or more of Georgia’s 159 counties and county health departments (47). The Atlanta metropolitan area is served by Cobb-Douglas, Fulton, Clayton, Gwinnet-Newton-Rockdale (GNR), and DeKalb public health districts. These public health districts contain the highest population densities and, besides

**Figure 1.**
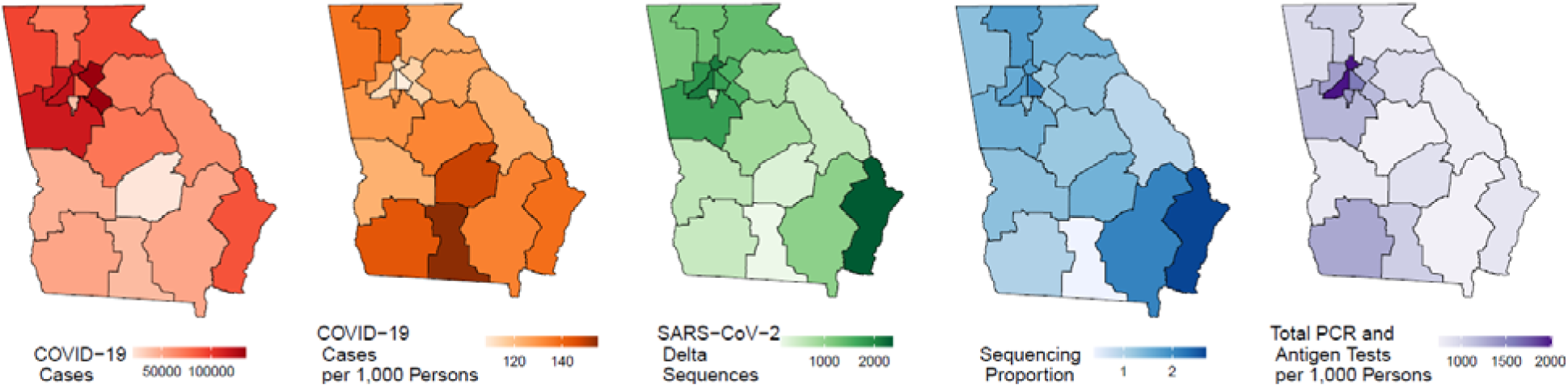
COVID-19 cases, sequences, and tests. More information about Georgia’s public health districts and detailed demographic statistics are in Supplementary Table 1.

Clayton, the largest population sizes in the state (Supplementary Table 1). The total number of PCR and antigen tests ranged from 751 to 2,007 per 1,000 persons; relatedly, the number of PCR-confirmed and antigen-positive COVID-19 cases reported during this study period ranged from 21,536 to 134,419 cases. Of these, the proportion sequenced ranged from 0.4 to 2.7% (Figure 1, Supplementary Table 1). While the highest case counts were in GNR, Fulton, and Cobb-Douglas, most sequences were collected from the Coastal public health district, which has the highest proportion of sequencing. COVID-19 incidence ranged from 109 to 155 cases per 1,000 persons. The highest COVID-19 incidence rates were found in South, South Central, and Southwest districts (Figure 1, Supplementary Table 1).

### Introductions into Georgia

Our final dataset consisted of 4,937 Georgia sequences and 4,846 representative worldwide sequences, including 6 continents, 122 countries, and 50 states, collected between August 1, 2020 and January 25, 2022. An initial time-scaled tree of 9,783 sequences was used to investigate introductions of the Delta variant into Georgia and onward viral migration between public health districts. We inferred 344 cumulative introductions into Georgia based on the ancestral state reconstruction of internal nodes, estimating the number of transitions from a non-

Georgia node to a Georgia node (n = 102) or to a Georgia tip, referred to as a singleton (n = 242). Of these 242 singletons, 155 resulted from distinct introductions, but 87 were nested within clusters. Singletons represent importation events associated with a single case; however, very low sequencing coverage implies that many of these singletons might actually represent small, undetected clusters. To gain insight into how long Delta was spreading in the state prior to detection in sequencing data after each introduction, we quantified the gap between the estimated time to most recent common ancestor for each introduction event and its earliest sampled sequence. We estimated about one month between introduction and detection (median = 25 days, days, IQR = 12-50 days), highlighting sparse sampling frequency (Figure 2). Singletons that were not nested within a cluster often stemmed from a polytomy. This indicates substantial uncertainty around the ancestor node, so we estimated the detection for these separately (median = 194, IQR = 74.5-373 days) (Supplementary Figure 4).

**Figure 2.**
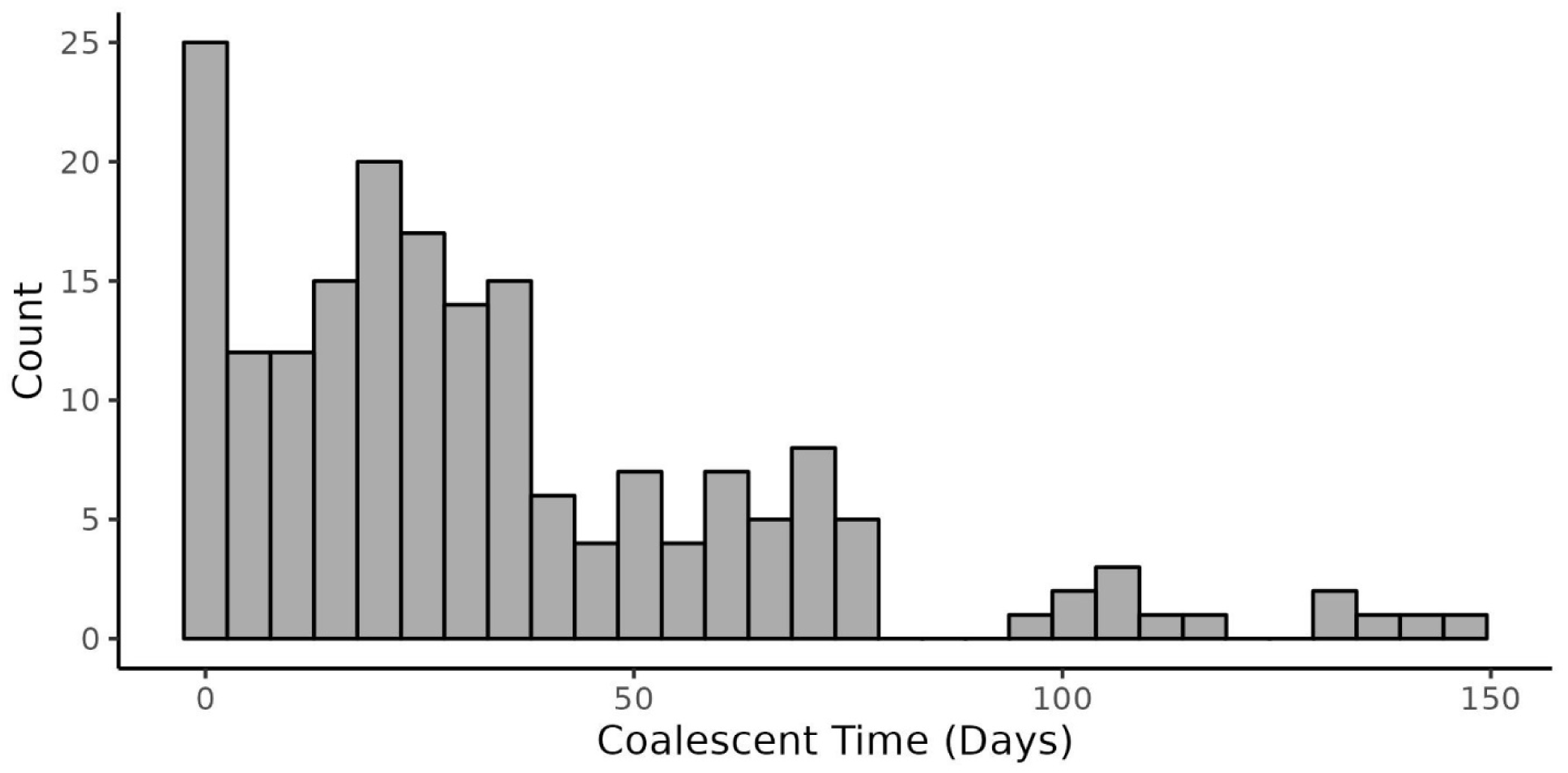
**Time to first detection of clusters after introduction into Georgia highlights sparse sampling frequency**. 102 distinct introduction events into Georgia resulted in at least 2 sequenced cases. Coalescent time between introduction event and earliest sequence was calculated to quantify the lag in detection time, which is indicative of sampling frequency.

For downstream analyses, clades (or “clusters”) descending from introduction events (n=102) were filtered to include those with at least three Georgia sequences (n=58) and sufficient branch support (n=34) (Supplementary Figure 5). These distinct clades represent Georgia transmission clusters, indicating at least 34 statistically supported, independent introductions of the Delta variant that successfully established transmission chains in Georgia. The median cluster size was 8 sequences (IQR: 4.3–19.5), with Georgia sequences comprising a median of 87.5% of each cluster (IQR: 77.3–100%). We inferred the TMRCA for each cluster and visualized the cluster sizes over time. We found that clusters became established in Georgia between April 2021 and November 2021 (Figure 3). The presence of many small clusters with sequences collected sporadically over an extended period indicates sparse sampling. Conversely, small clusters with sequences collected over a short time frame likely reflect limited local transmission. Several medium and large clusters with more normally distributed sequences collected throughout the epidemic (e.g., Cluster 9 and 34) indicated substantial local transmission and dense sampling.

**Figure 3.**
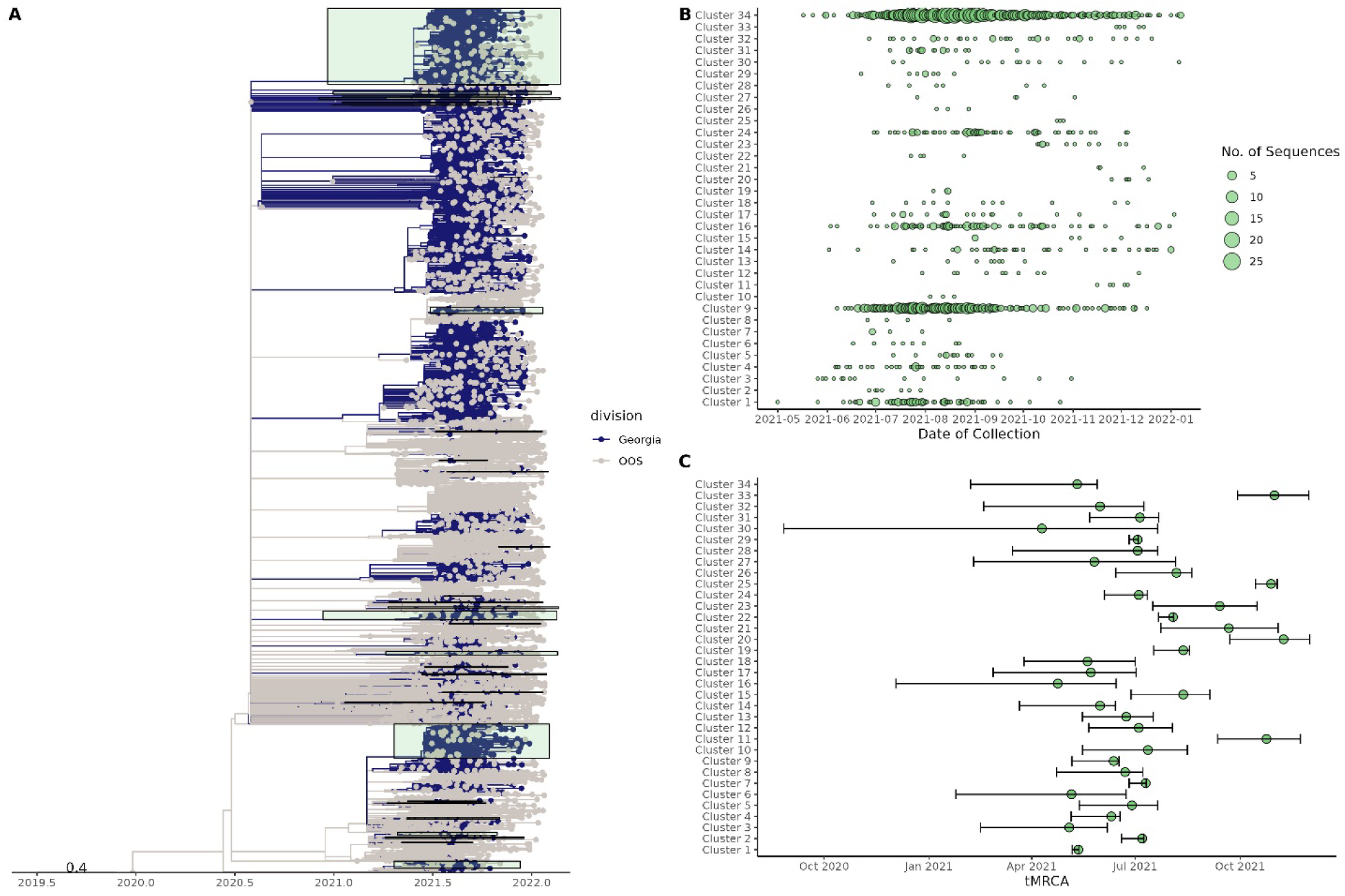
Characterization of Georgia clusters (n=34). (A) Internal nodes and tips are colored based on their inferred location or site of collection, respectively, either in Georgia (blue) or out of state (OOS) (gray). Identified clusters are highlighted in green. (B) The number of sequences over time, demonstrating large clusters like Cluster 9 and Cluster 34 that are likely more densely sampled compared to smaller, more sporadic clusters. (C) Date of the most recent common ancestor for each cluster estimated in original ML tree.

Highly consistent results for the number of inferred introduction events, number of clusters, and cluster sizes were obtained after running analyses on four additional resampled datasets suggesting our observed results were robust to resampling (Supplemental Table 2).

### Source-Sink dynamics in Georgia

To provide a more robust ancestral state reconstruction of local transmission in Georgia, Bayesian phylogeographic analyses were performed on clusters, using public health district. Clusters that consisted of fewer than 5 sequences or were homogeneous on public health district were excluded, resulting in 24 clusters (Supplementary Figure 6). We estimated absolute metrics, including all transitions (Markov jumps) between states along phylogenetic branches, averaged over MCMC states. We also measured the time (Markov rewards) spent in each state between transitions, represented as the average total time over MCMC states. Markov rewards were visualized for all clusters to inspect the extent of inter-cluster variation (Supplementary Figure 7). In addition, Markov jumps were visualized for small clusters (<10 sequences) (Supplementary Figure 8) and medium clusters (10 – 99 sequences) (Supplementary Figure 9). However, downstream analyses focused on Cluster 9 (n = 388 taxa) and Cluster 34 (n = 863 taxa) because they were large densely sampled clusters that spanned the entire study period and were inclusive of all possible discrete traits.

In Cluster 9, the highest average number of transitions were inferred from Coastal to out of state (n = 85), followed by out of state to District 4 (n = 34) and Coastal to Southeast (n = 22) (Supplementary Figure 10). Upon examining these transitions over time, we found that transitions were concentrated in Summer 2021 (Supplementary Figure 11). Markov rewards estimated that viruses circulated the longest in Coastal (33%), out of state (20%), and District 4 (11%) (Supplementary Figure 7). Beyond these absolute statistics, we also estimated the relative risk of each public health district as a source of transmission for a specific sink and assessed its potential for enhanced risk (compared to other sources). We found that the Coastal district was statistically a more likely source for out of state transitions compared to all other possible sinks (RR = 5.58, 95% HDI: 1.81, 11.99) (Supplementary Figure 12). While this is the only source with significant relative risk for transitions to out of state locations, the confidence intervals among the other source districts overlap, suggesting that their contributions to the sink were not significantly different from one another. Furthermore, although Coastal is a significantly more likely source for the Southeast district compared to other sinks (RR = 2.45, 95% HDI: 1.46, 3.40), there appears to be little heterogeneity among sources, suggesting no significant differences in their contributions to the Southeast district. We found no evidence that the relative risk from out-of-state to District 4 is significant.

In Cluster 34, the highest average number of transitions occurred from out of state to Fulton (n = 92), Fulton to District 4 (n = 91), and Fulton to Cobb-Douglas (n = 70) (Supplementary Figure 10). These transitions are concentrated in summer 2021 (Supplementary Figure 13). Markov rewards revealed the importance of Fulton (29%), out of state (20%), and District 4 (14%) in seeding infections (Supplementary Figure 7). Our relative risk analysis suggests that out-of-state locations are a more likely source for Fulton compared to all other possible sinks (RR = 39.39, 95% HDI: 0.14, 117.96) (Supplementary Figure 14). However, this finding was not statistically significant. While three other sources (South Central, South, and Clayton) have significant but minimal effect sizes, the wide HDI interval indicates substantial uncertainty in the contribution of out-of-state locations to Fulton. We found no significant evidence of relative risk from Fulton to District 4 or to Cobb-Douglas.

### Migration dynamics in Georgia

We assessed the importance of individual transitions to reconstruct the transmission network across the state. We applied a single rate matrix to jointly estimate transition rates across all clusters. This approach adjusts for inter-variation of cluster makeup and size. Out of 342 transition rates, we identified 34 decisive rates (BF > 100) that were statistically supported (Posterior Probability (PP) > 50%). Of these, the highest rates were Fulton to District 4 (4.3 transitions/year, PP = 97.7%), DeKalb to out of state (3.8 transitions per year, PP = 99.5%), and Fulton to DeKalb (2.8 transitions/year, PP = 88.3%) (Figure 4).

**Figure 4.**
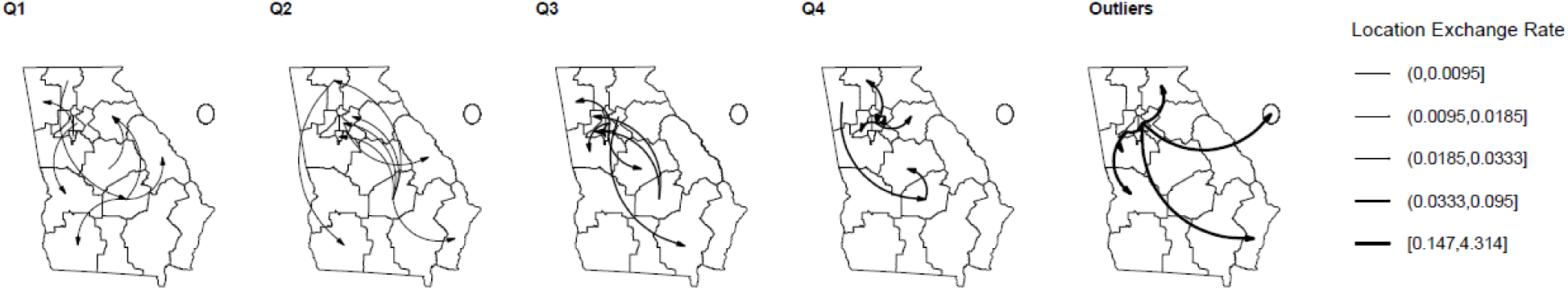
BSSVS transition rates among public health districts and out of state locations, by quantile. Decisive (BF > 100) and statistically supported rates (Posterior Probability (PP) > 50%) are divided by rate quantile (Q1-Q4). Outliers (outside of 1.5x IQR) were removed prior to calculating transition rate quantiles and are presented separately to highlight particularly high transition rates (e.g., Fulton to District 4, DeKalb to out of state, and Fulton to DeKalb). The circle denotes out of state locations.

This joint estimation approach assumes that the underlying mechanism of spatial diffusion remains consistent across clusters. To assess concordance with individual analyses, we examined individual clusters that encompassed all possible discrete traits. Of the 34 decisively supported transition rates that were identified in the joint analysis, 22 were also decisively supported in the individual clusters analyses. These included seven of the nine decisively supported transition rates in Cluster 9: GNR to Southeast; North Georgia to Southwest and South Central; South Central to Fulton, Coastal, Clayton, and East Central. Similarly, 13 of the 16 decisively supported transition rates in Cluster 34 were also identified in the joint analysis: Northwest to South Central; South Central to DeKalb; and DeKalb to out of state, District 4, Fulton, East Central, West Central, North Central, Coastal, North Georgia, Northeast, Northwest, and Clayton (Supplementary Figure 15). Despite differences in the number of taxa and distribution of public health districts among individual clusters, the results remained highly concordant with the joint analysis. However, even though Clusters 9 and 34 were by far the largest, the joint analysis captured a more comprehensive representation of spatial diffusion across clusters.

Lastly, we evaluated the robustness of our jointly inferred transmission network upon rerunning our entire pipeline across additional resampled datasets. Across our 5 replicates, we identified between 17 – 34 decisive transition rates that were statistically supported (Supplementary Figures 16-17). Of these, we found 10 decisive, statistically supported common rates across all five subsamples: North Georgia to South Central and South Central to: Clayton, Coastal, DeKalb, East Central, Fulton, GNR, North Central, North Georgia, and Northeast (Figure 5).

**Figure 5.**
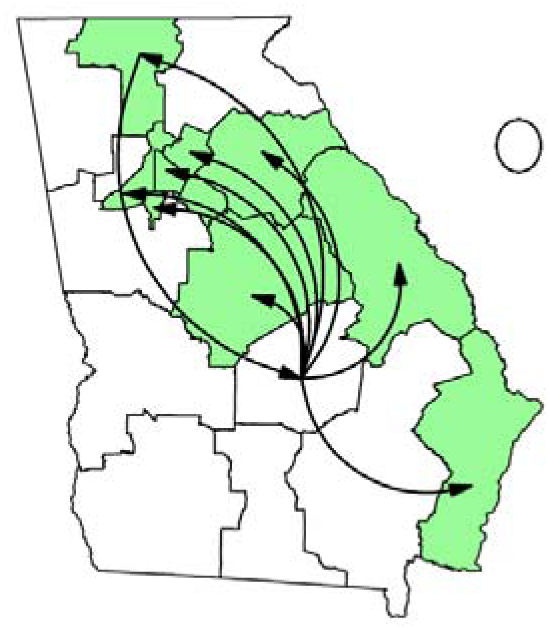
Decisive and statistically supported transition rates across five resampled datasets. Rates inferred from a joint likelihood Bayesian framework are decisive (BF > 100), statistically supported (PP > 50%), and robust across five resampled datasets. The circle denotes out of state locations.

These rates are robust and not sensitive to the subsample, likely capturing the underlying connectivity across the state during the Delta wave.

## Discussion

In this study, we estimated the number and timing of SARS-CoV-2 Delta introductions into Georgia, described transmission heterogeneity, identified key sources and sinks of transmission, and inferred the statewide transmission network during the Delta wave. We identified a one-month lag time between introduction events and first detection. In addition, we found that many small clusters were comprised of sequences sporadically collected over a long timeframe. Taken together with low sequencing rates across public health districts, our study highlights generally sparse sampling and/or sequencing frequency. While the average size of clusters was quite small, a few introduction events led to large and sustained circulation in Georgia. Given heterogeneity in the size, sampling, and distribution of public health districts, we jointly inferred transmission rates across all clusters to describe overall statewide epidemic dynamics. We found overwhelming and robust evidence of the role of South Central as a key source in the COVID-19 epidemic in Georgia.

We built a rapid pipeline with several advantages compared to previous studies. The highest number of cases are in public health districts that make up the Atlanta Metropolitan Area, but most sequences come from the Coastal public health district. In order to adjust for this sequencing bias, we subsampled sequences proportionately to the number of reported cases in each public health district. As these inferences may guide public health decision-making, we added node support values to the preliminary maximum likelihood phylogenetic tree and restricted clusters to those with very high support. Since standard bootstrapping to assign node support has become prohibitively computationally expensive for large phylogenies, we applied ultrafast bootstrap and single branch support tests. This allowed us to incorporate phylogenetic uncertainty and avoid information loss and bias that comes from using a single tree estimate (9). Compared to standard bootstrap support (SBS), UFBoot2 is nearly 800x faster and has been shown to produce more unbiased branch support values (29). Similarly, rapid single branch support methods, such as aBayes and SH-aLRT, have demonstrated accuracy, power, and robustness in empirical and simulated studies (32). Finally, we changed the starting seed at the subsampling step, rerunning our entire pipeline from alignment to joint estimation. This allowed us to test if our inferences are robust to resampling in addition to model stochasticity.

We detected at least 344 introductions into Georgia from an analysis of 9,783 Delta sequences collected between August 1, 2020 and January 25, 2022. The state of Georgia has extensive national and international connectivity facilitated by Hartsfield-Jackson International Airport, one of the world’s busiest air travel hubs. While the inferred number of introductions is certainly an underestimate due to low sequencing proportions, it implies that a small proportion of the sequences in this analysis were attributed to novel introduction compared to local transmission. While the median cluster size in our study was quite small, we found that a few introductions resulted in a large number of sequences. This is consistent with studies that have reported transmission heterogeneity of SARS-CoV-2 (11). For example, Lemieux et al. 2021 described two superspreader events, one of which was contained in a skilled nursing facility, while the other was exported and resulted in substantial regional, national, and international spread (14). Similarly, a phylogenetic study of SARS-CoV-2 sequences in Louisiana found that a single introduction following Mardi Gras events was responsible for the majority of early transmission within the state (15).

Importantly, we estimated that clusters were circulating in Georgia for about one month on average between the introduction event and first detection, highlighting generally sparse sampling frequency. This is consistent with a previous phylogenetic study in Georgia that found that SARS-CoV-2 was circulating within Georgia for at least two to four weeks before the first detected infection was reported to the GA DPH (10). In our study, sparse sampling can be visualized by the presence of many small clusters with sequences sporadically collected over a long timeframe. On the other hand, a few small clusters with sequences collected in a short time frame are indicative of true limited local transmission. In addition, we found several medium and large clusters that had sequences that were more normally distributed over the entire time period of the epidemic in Georgia, suggesting dense sampling and substantial local transmission.

Markov jumps provide insight on the intensity of transitions between states, while relative risk specifically assesses the enhanced risk of a public health district as a viral source. We found strong evidence of many public health districts to be more likely sources for particular public health districts compared to other sinks. Yet, overlapping confidence intervals suggested that no particular source’s contribution to a sink was statistically different from others. Small sample sizes in the number of introductions to certain locations may mean that we have reduced statistical power to detect relative risk. The HPD intervals for some of the relative risk measures are also very wide, indicating statistical uncertainty, likely due to low numbers of introductions. Relative risks with these wide confidence intervals should be interpreted with caution.

BSSVS rates assess the structure of the network by identifying connections that contribute the most to the transmission process. The joint analysis represents a more comprehensive picture of spatial diffusion across the state, assuming an underlying statewide connectivity that individual clusters may not fully capture. Even large clusters that include all public health districts may represent different outbreaks across the state, each with distinct transmission dynamics. Despite variations in cluster size and district distribution, the rates observed within each cluster were highly concordant with those from the joint analysis.

Importantly, we found robust, overwhelming evidence for the role of the South Central public health district as a key source of Delta transmission to the rest of the state, including four of the five Atlanta districts. These rates are robust and not sensitive to the subsample, likely capturing the underlying connectivity across the state during the Delta wave. We also found that the highest incidence rates were in South, South Central, and Southwest public health districts. Taken together, this consistent with findings from the early months of the COVID-19 pandemic in Georgia. Dougherty County, in the rural Southwest public health district, was reported as the first epicenter of COVID-19 in the state (48,49) following a superspreading event at a funeral. Moore et al. 2020 also found that southwestern rural Georgia had the highest increases in COVID-19 incidence rates from March – April 2020. Counties with the highest mortality rates had a higher proportion of adults aged 60+, adults earning less than $20,000 annually, and residents living in rural communities, compared to counties with lower mortality rates. These counties also had very limited healthcare resources, including intensive care unit beds and primary care physicians (50).

Southwest counties identified as COVID-19 hot spots for prevalence and mortality have been historically associated as hot spots for other diseases, such as stroke and sepsis, suggesting continuing disparities in health outcomes (50–52). Populations in South Georgia have a disproportionate burden of cardiovascular disease, obesity, diabetes, and other co-morbidities that are risk factors for infections. These conditions make a population more vulnerable to COVID-19 infection, severe disease, and mortality (48). Identifying geographic areas that have the highest COVID-19 burden allows targeted resource allocation by policy makers, public health professionals, and critical care providers. This may be important in preparing for future SARS-CoV-2 waves for at-risk populations. As the virus spread, southern Georgia may have experienced a disproportionate number of COVID-19 cases due to social (non-medical) determinants of health that can influence health outcomes (48). Future studies could benefit from the addition of non-medical factors in models, such as socioeconomic status, healthcare access, race/ethnicity, age, and urban/rural status. These variables could provide deeper insights into transmission drivers, particularly when coupled with contact tracing efforts to generate more comprehensive datasets.

We attempted to correct for several anticipated limitations that are inherent to genomic surveillance, such as sequence quality and sampling bias. Lags in sequence submission to public repositories, low sampling and sequencing rates, and missing metadata present challenges for robust epidemiological inferences of circulating and emerging variants and epidemic dynamics. Markov jumps and rewards are sensitive to sample sizes, so it is possible that our inferences for source-sink dynamics for individual clusters are biased towards public health districts that are overrepresented in each cluster, despite our efforts to control for this impact. Additionally, low sampling and sequencing rates likely resulted in underestimates of total introductions into Georgia (if neither index cases nor descendants are sampled), underestimates of sublineage sizes (if not all descendants are sampled), and overestimates of the proportion of singletons (which may have resulted from unsampled transmission chains) (22).

Our study addresses a critical need for methodologies and data-driven recommendations for public health practice, particularly given large, dynamic, and integrated datasets. These inferences can inform targeted outbreak investigation, as well as guide dissemination of public health resources (e.g., contact tracing, mobile testing units). In addition, our pipeline can be adapted for future outbreaks. Academic-public health partnerships are an important resource for public health research; through our collaborations we have been able to link genomic sequences with spatial metadata to answer important questions about COVID-19 spread in Georgia, including the contribution of importations to local spread, key sources and sinks of transmission, and lag time to detect out of state introductions.

## Data Availability

All R scripts are available at https://github.com/Gabriella-Veytsel/Clusters. Genome sequences and associated metadata are published in GISAID’s EpiCov database, available at https://doi.org/10.55876/gis8.251125to, https://doi.org/10.55876/gis8.251124px, https://doi.org/10.55876/gis8.251124no and https://doi.org/10.55876/gis8.251124bd.

## Data Availability

All R scripts are available at https://github.com/Gabriella-Veytsel/Clusters. Genome sequences and associated metadata are published in GISAID EpiCov database.

https://doi.org/10.55876/gis8.251125to

https://doi.org/10.55876/gis8.251124px

https://doi.org/10.55876/gis8.251124no

https://doi.org/10.55876/gis8.251124bd

## Acknowledgments

We thank the Georgia Department of Public Health epidemiologists, laboratorians and bioinformaticians. We gratefully acknowledge all data contributors, i.e., the Authors and their Originating laboratories responsible for obtaining the specimens, and their Submitting laboratories for generating the genetic sequence and metadata and sharing via the GISAID Initiative, on which this research is based.

## Author Contributions

G.V., L.L., G.S., and J.B. conceptualized the study. G.V. conducted data analysis, data interpretation, data visualization, and manuscript writing. H.D. contributed resources. J.B. and L.C. provided supervision and acquired financial support. All authors contributed to reviewing and editing.

---

**Supplemental Figure 1.**
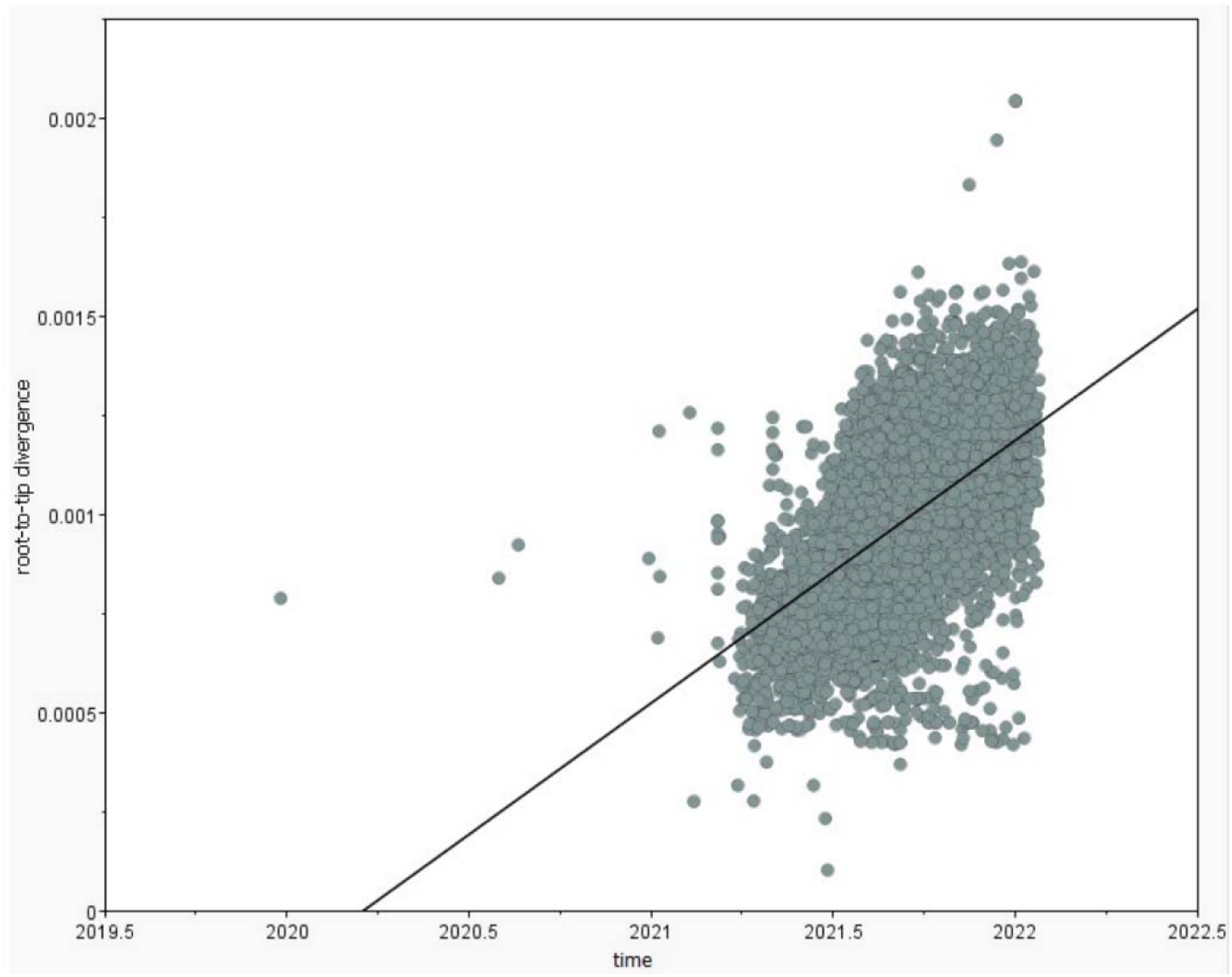
Tempest root-to-tip (RTT) regression plot.

**Supplementary Figure 2.**
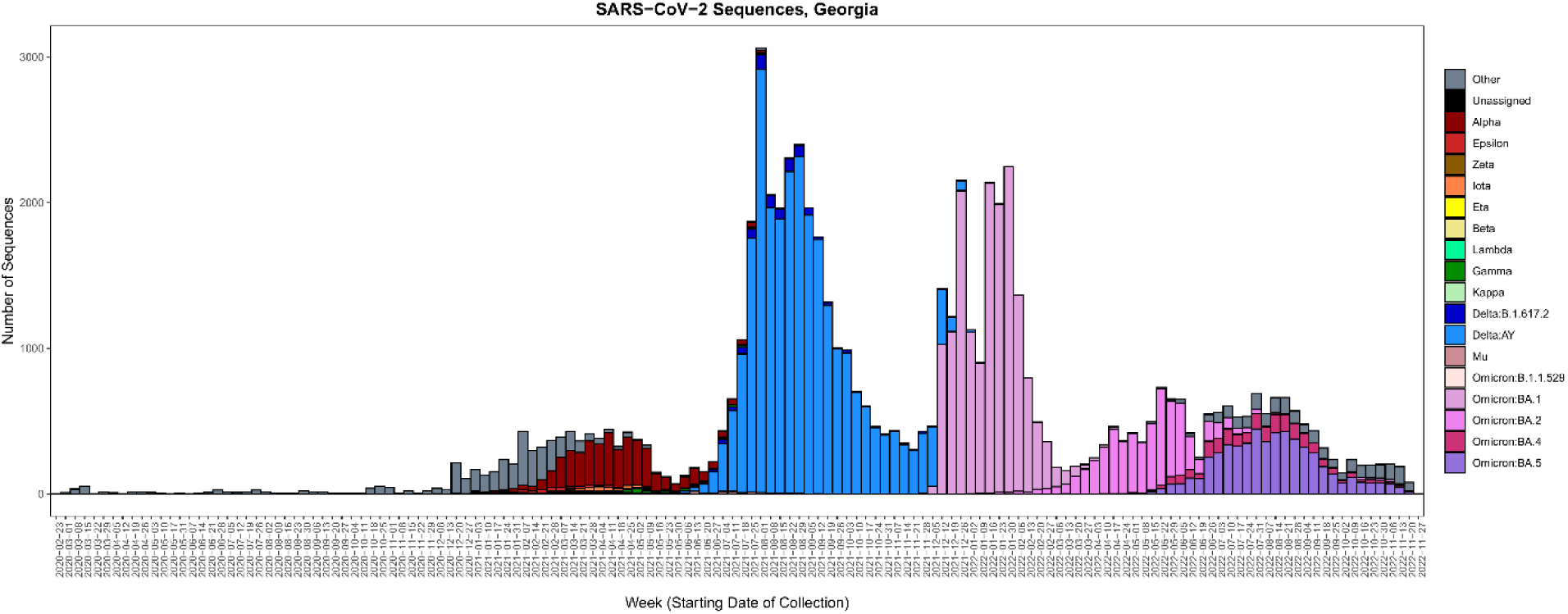
SARS-CoV-2 Delta Wave. All SARS-CoV-2 complete genomes collected in Georgia available in GISAID were downloaded on December 7, 2022, filtering sequences for complete collection date (n = 66,331). This dataset only served to define the Delta wave in context with other circulating variants. In Georgia, the first Delta sequence was collected on April 9, 2021 and the last on March 10, 2022. The Delta variant was the dominant circulating SARS-CoV-2 variant for 24 weeks (June 27, 2021 – December 11, 2021). Sequences available on GISAID (EPI_SET_251124bd, EPI_SET_251124no).

**Supplementary Figure 3.**
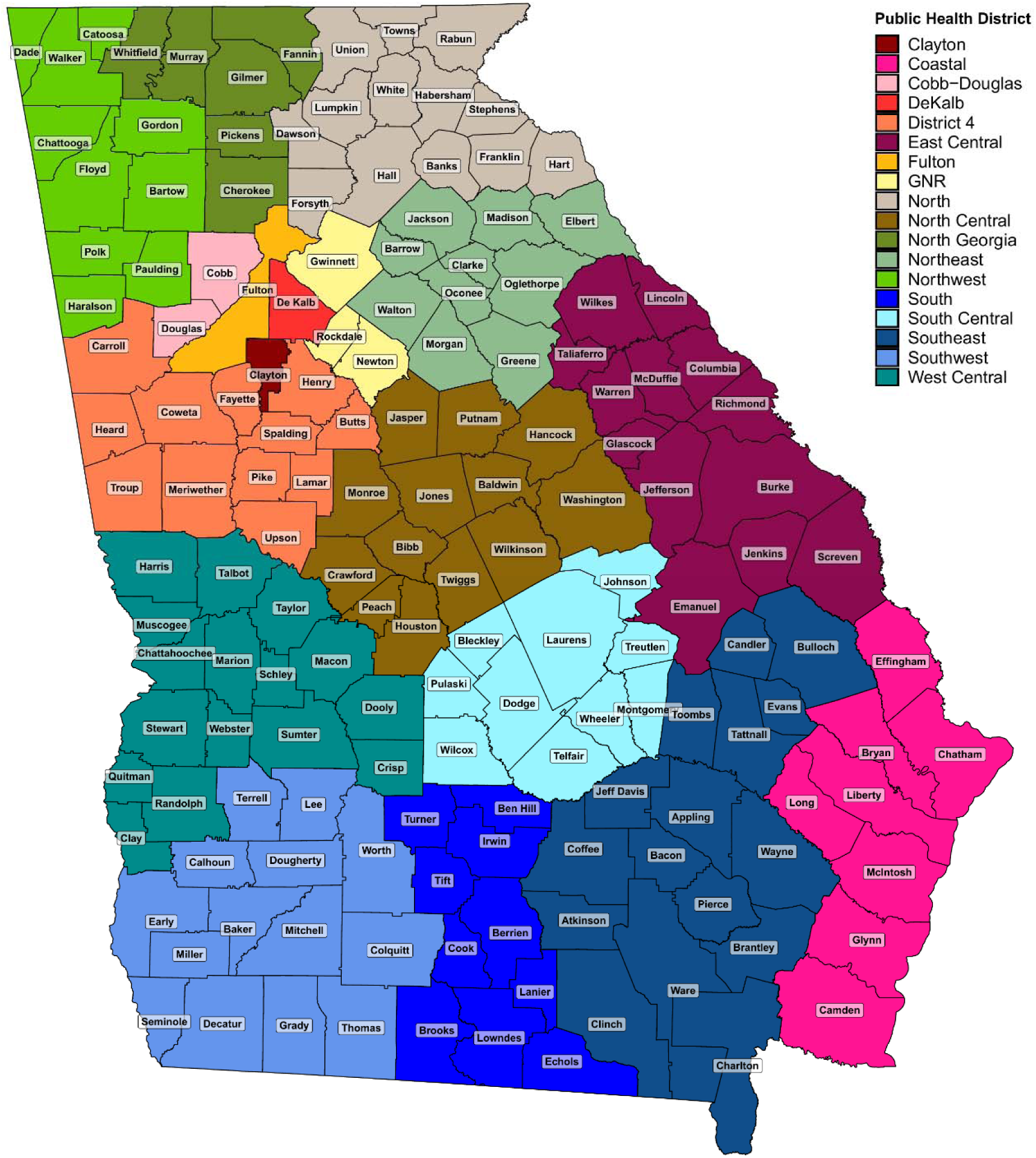
Map of counties and public health districts, Georgia.

**Supplementary Figure 4.**
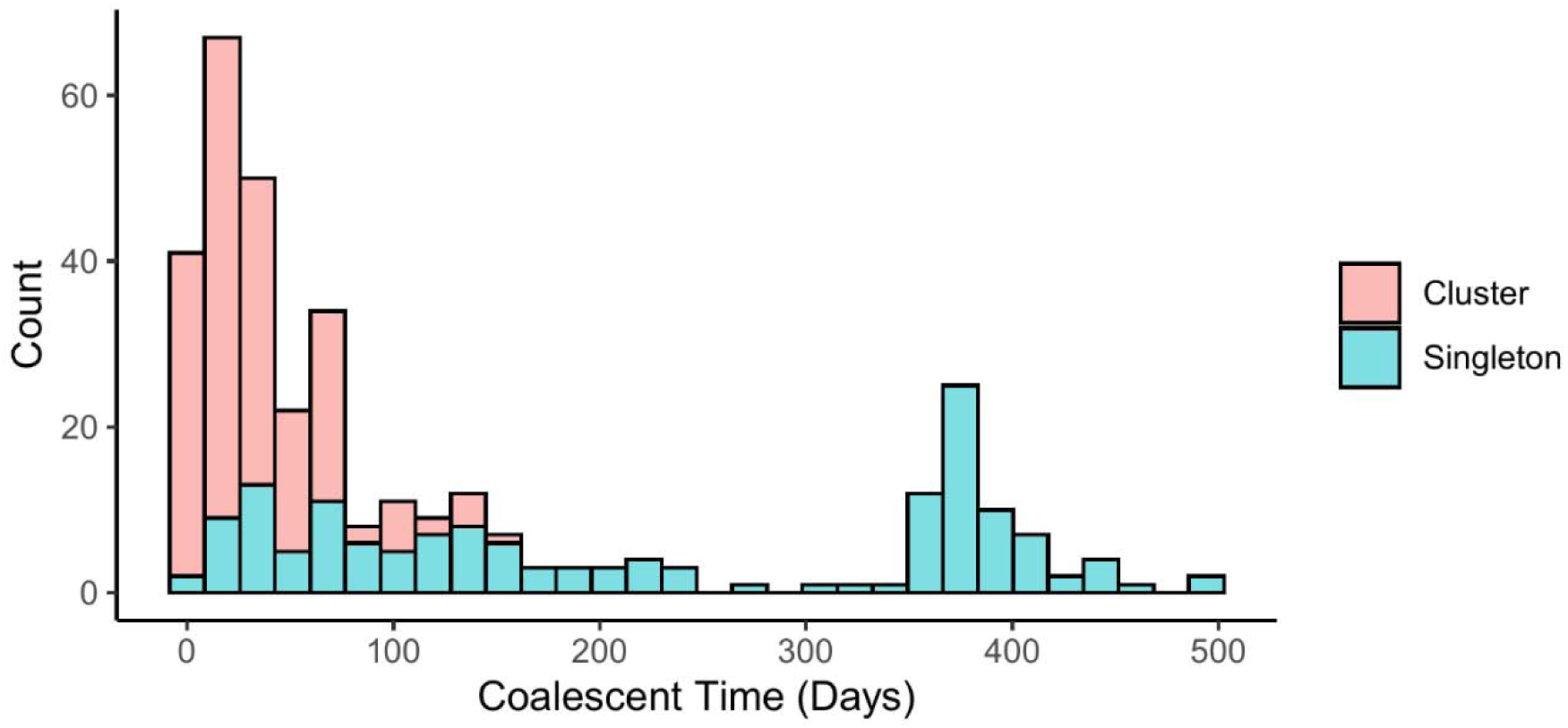
Time to first detection of clusters after introduction into Georgia inferred from maximum likelihood phylogenetic analysis. Time between introduction event (node) and first sequence detection in Georgia (earliest tip). Red indicates sequences that are associated with clusters, while blue indicates singletons that are not associated with a cluster. Low sequencing coverage implies that many of these singletons might actually represent small, undetected clusters. Sequences associated with a cluster have a lower time between introduction and detection, compared to sequences that are not. Singletons with a long time to detection stem from polytomies that form the spine of the tree. This indicates important uncertainty around the ancestor.

**Supplementary Figure 5.**
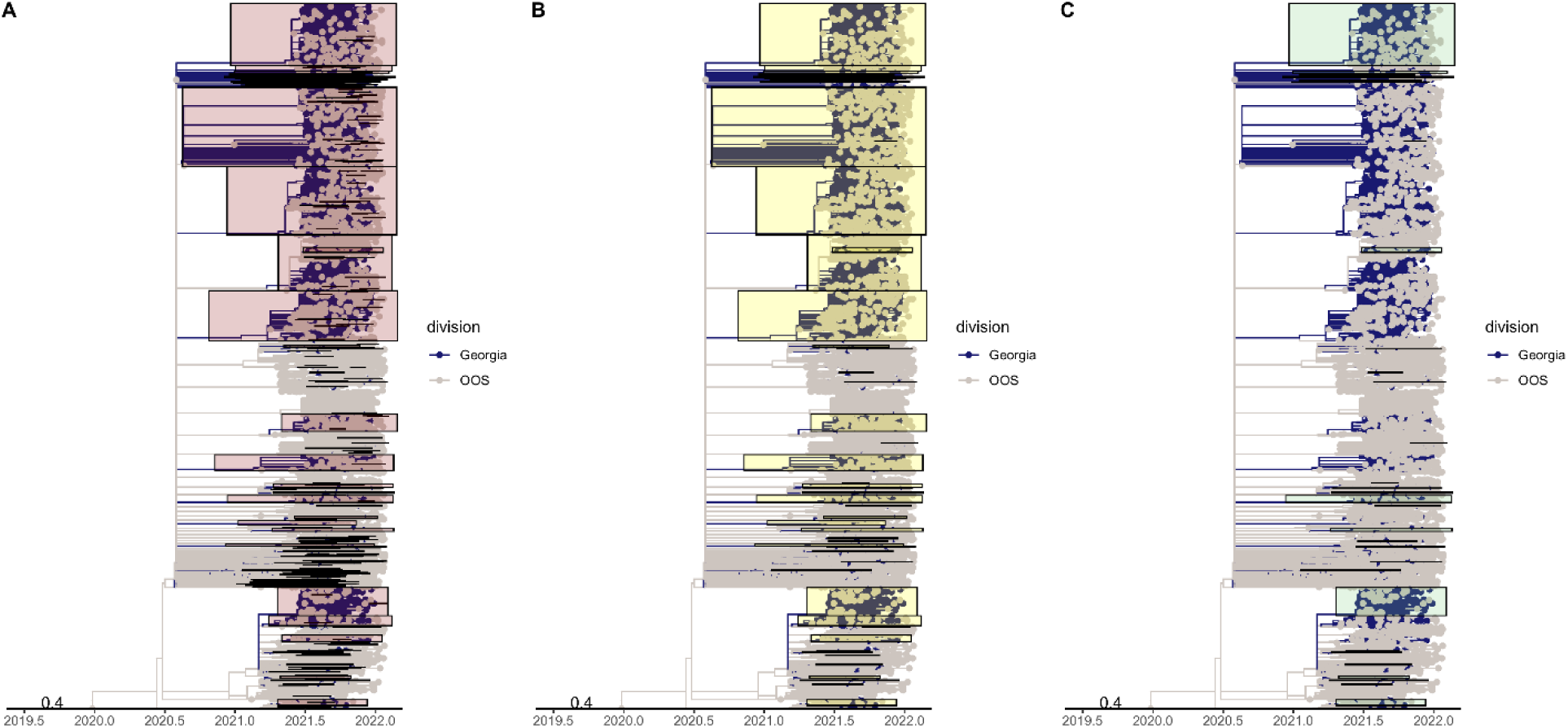
Initial Maximum Likelihood phylogenetic trees to identify Georgia clusters. (A) Clades descending from inferred introduction events are highlighted in pink (n=344). (B) Clades with >= 3 Georgia sequences are highlighted in yellow (n=58). (C) Statistically supported clades (or “clusters”) with >= 3 Georgia sequences (n=34), highlighted in green.

**Supplementary Figure 6.**
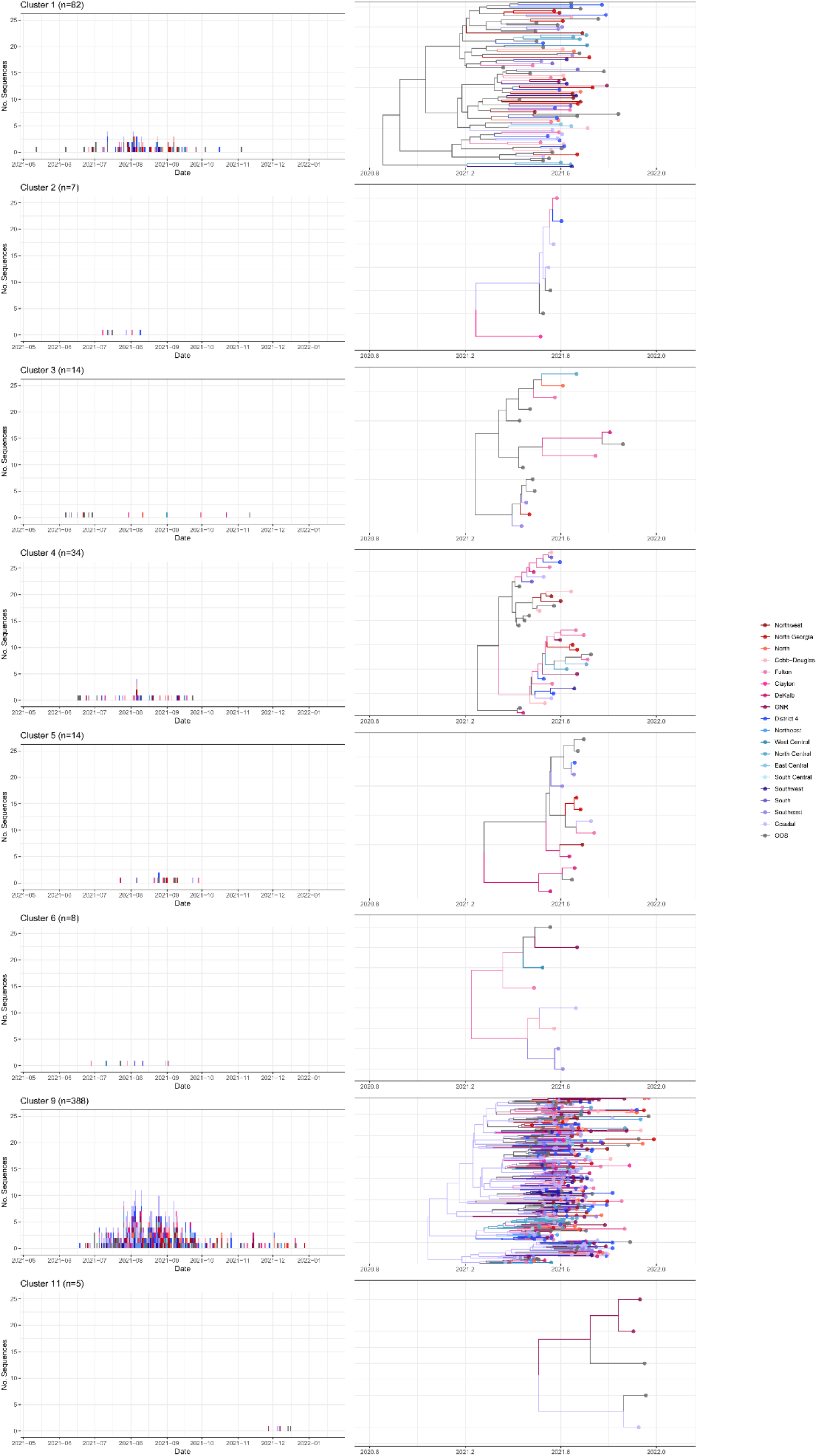
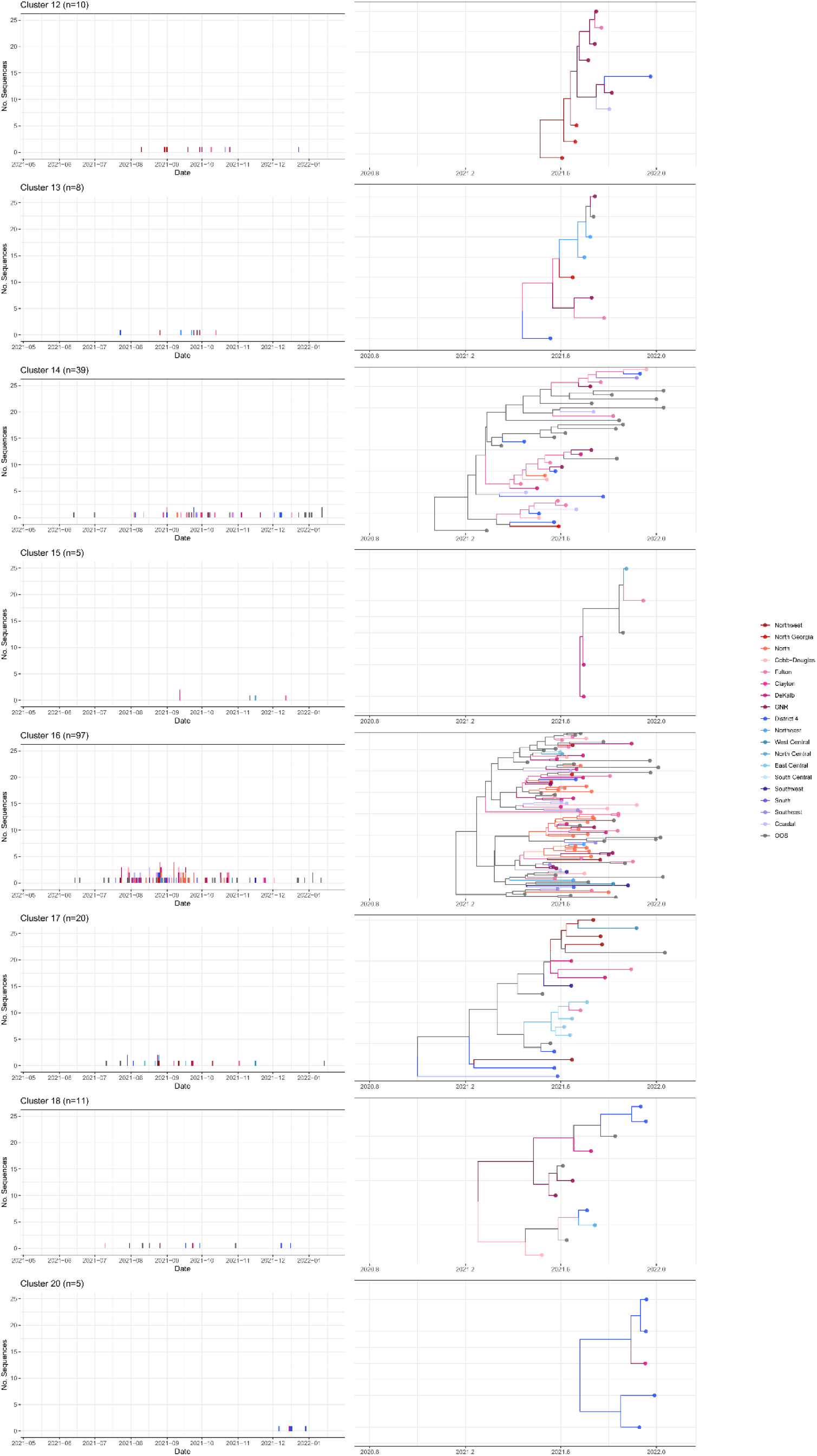
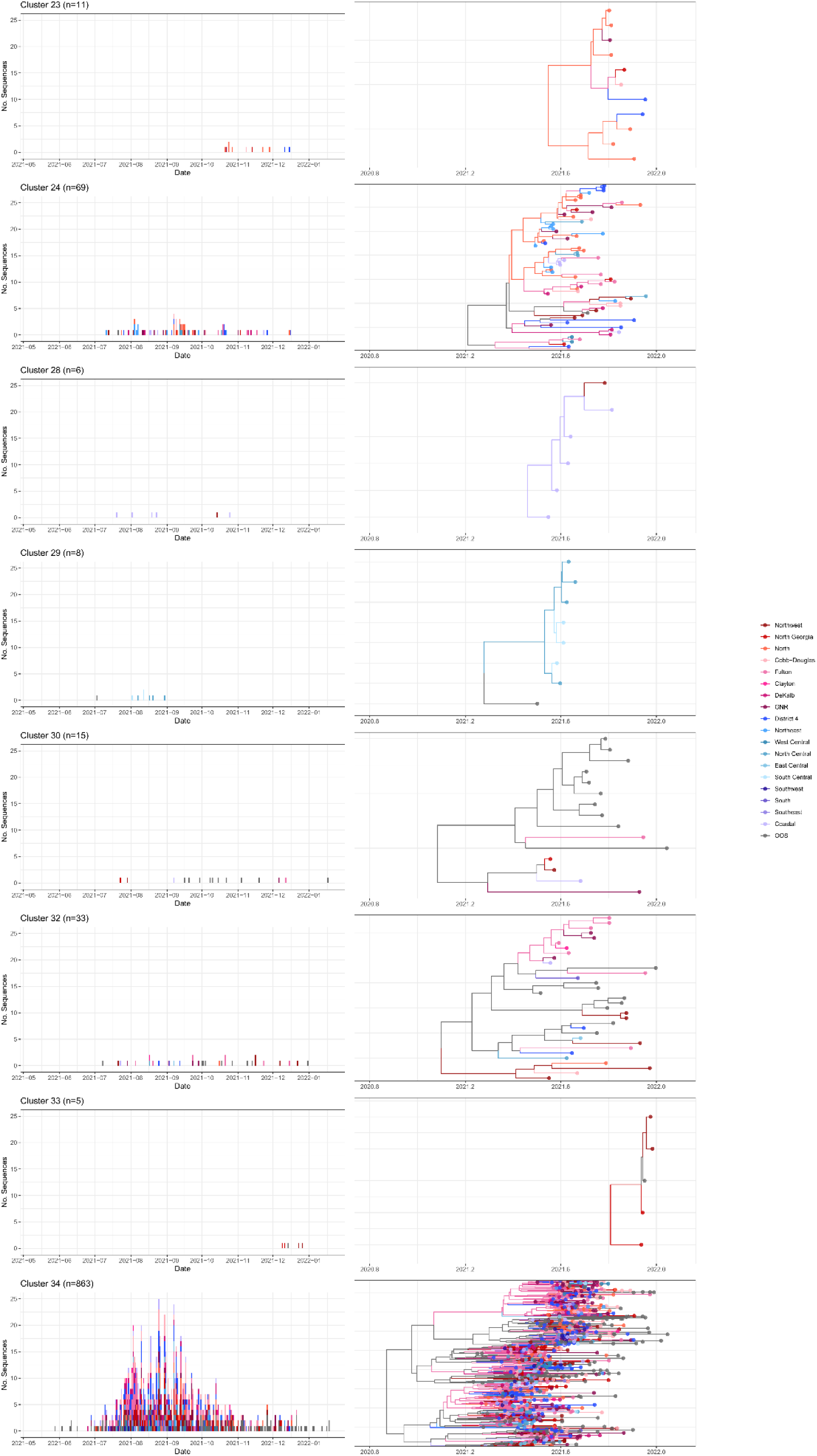
Distribution of sequences counts and Bayesian maximum clade credibility trees for each cluster (n=24). The bigger, more normally distributed clusters span the time period of the epidemic in the state and are likely more densely sampled than the clusters that are sporadic and long.

**Supplementary Figure 7.**
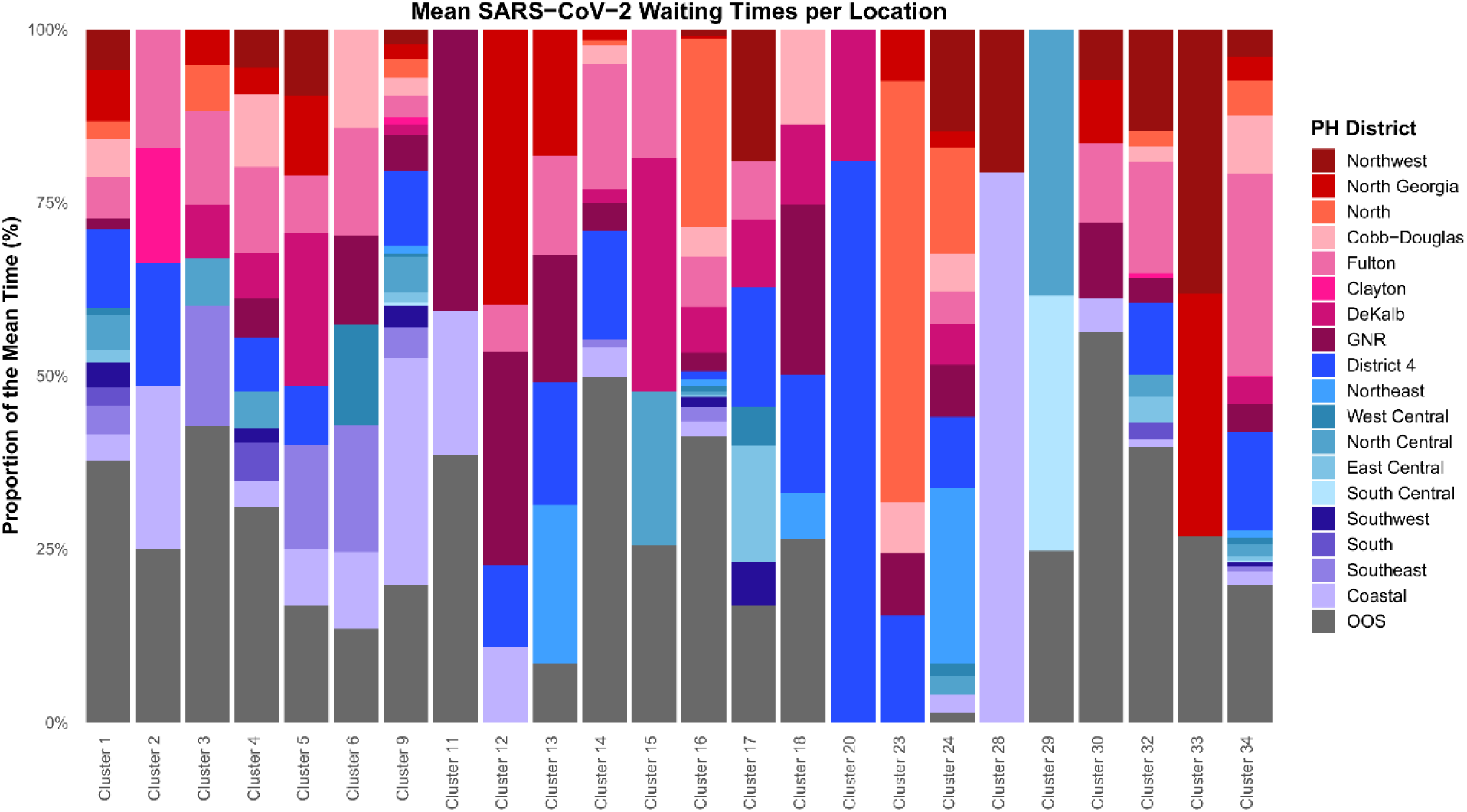
Markov rewards for individual clusters. Markov rewards for public health district show the waiting time for a given district across branches of the phylogeny over time.

**Supplementary Figure 8.**
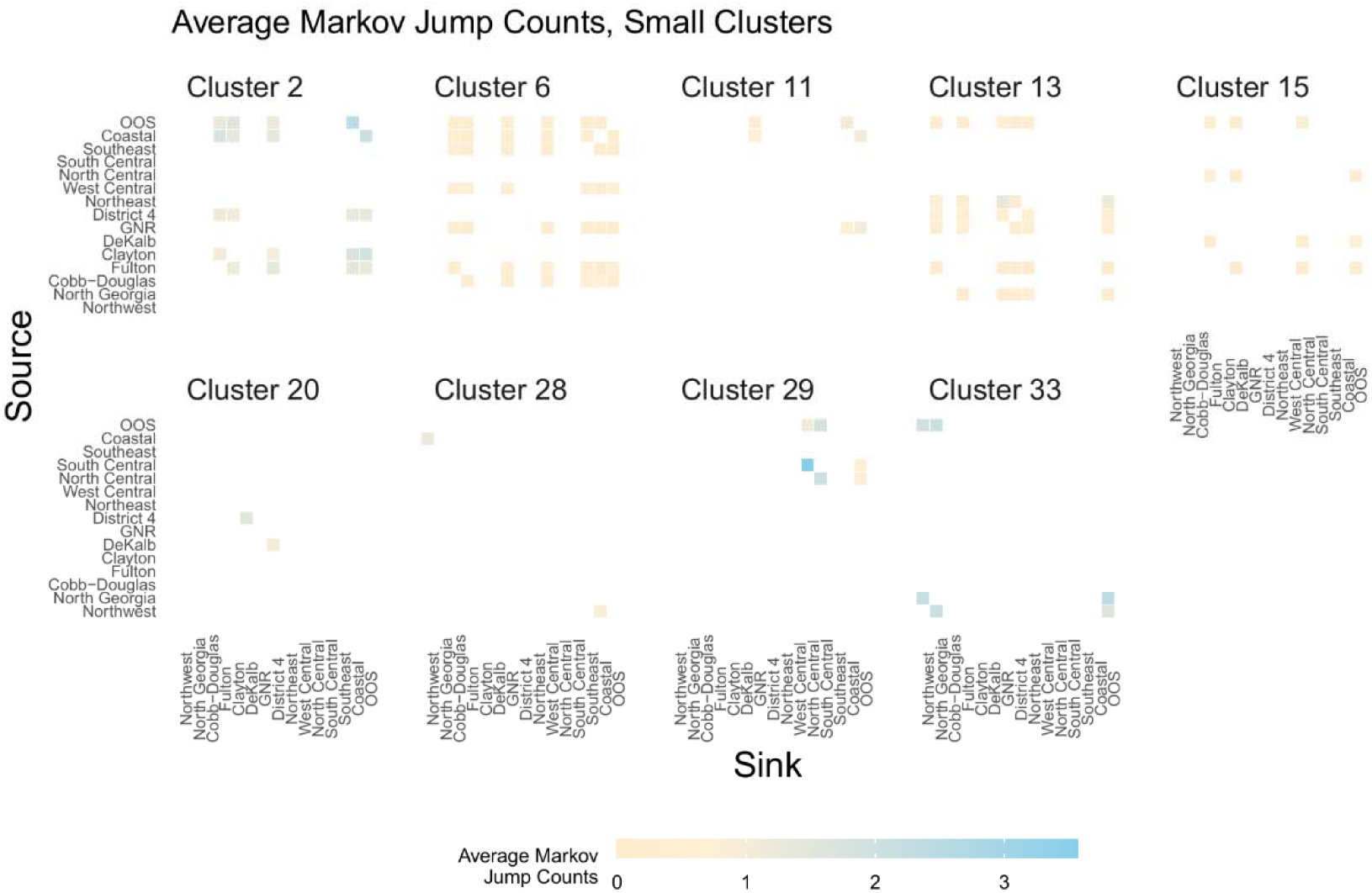
Markov jumps for individual, small clusters. Phylogenetic discrete trait analyses inferred transitions (Markov jumps) between public health districts over time along phylogenetic branches. Darkness of each cell is proportional to the absolute number of transitions from the first column to the second column over time.

**Supplementary Figure 9.**
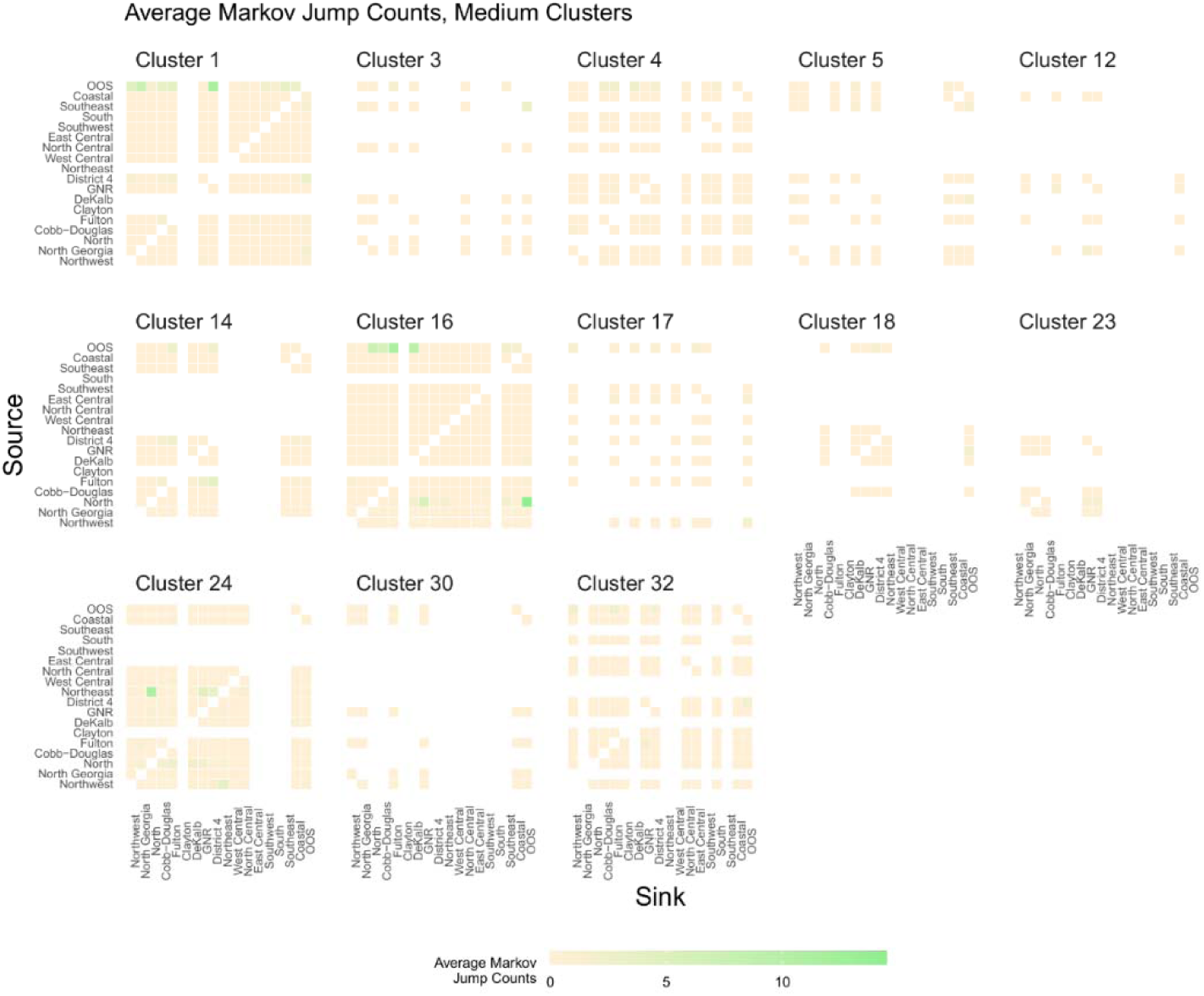
Markov jumps for individual, medium clusters. Phylogenetic discrete trait analyses inferred transitions (Markov jumps) between public health districts over time along phylogenetic branches. Darkness of each cell is proportional to the absolute number of transitions from the first column to the second column over time.

**Supplementary Figure 10.**
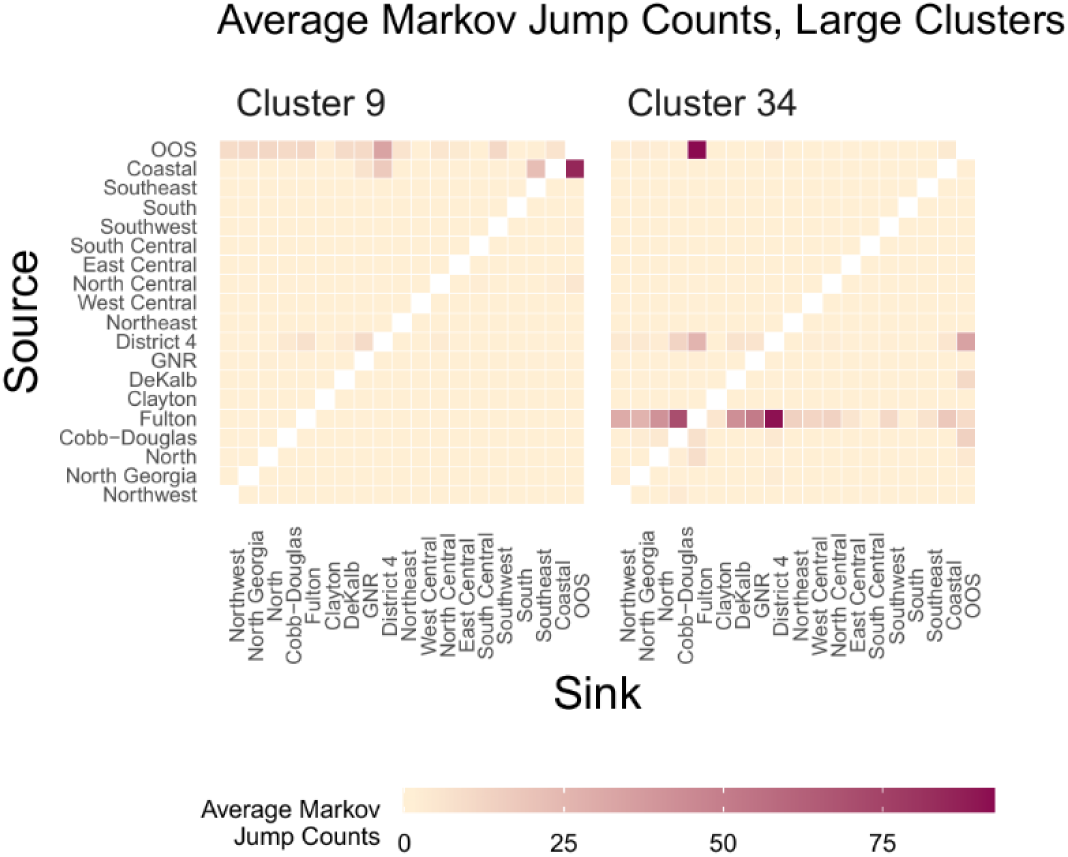
Markov jumps for individual, large clusters. Phylogenetic discrete trait analyses inferred transitions (Markov jumps) between public health districts along phylogenetic branches. Darkness of each cell is proportional to the absolute number of transitions from the first column to the second column over time.

**Supplementary Figure 11.**
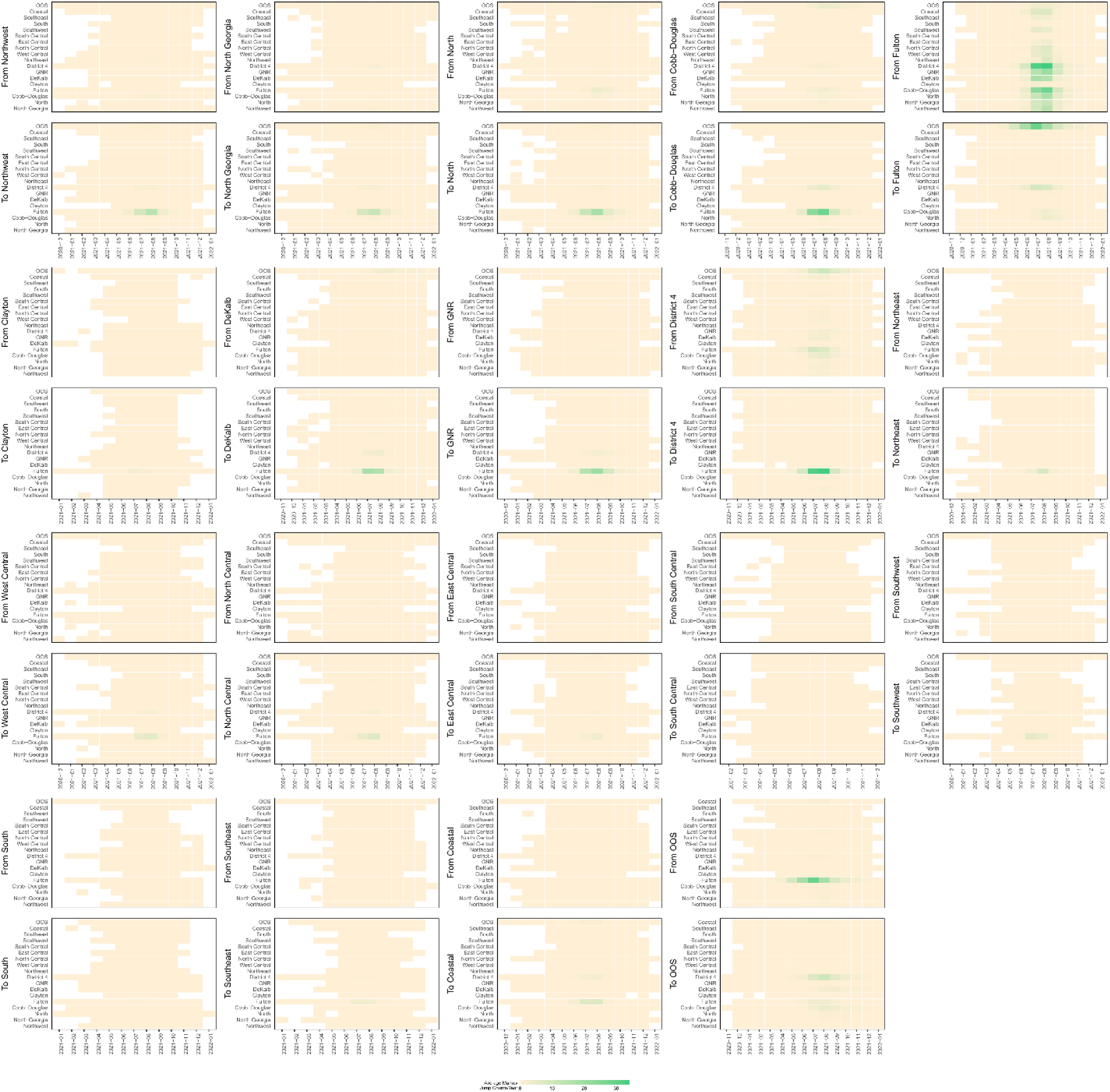
Markov jumps over time, Cluster 9. Phylogenetic discrete trait analyses inferred transitions (Markov jumps) between public health districts over time along phylogenetic branches. Darkness of each cell is proportional to the absolute number of transitions from the first column to the second column over time.

**Supplementary Figure 12.**
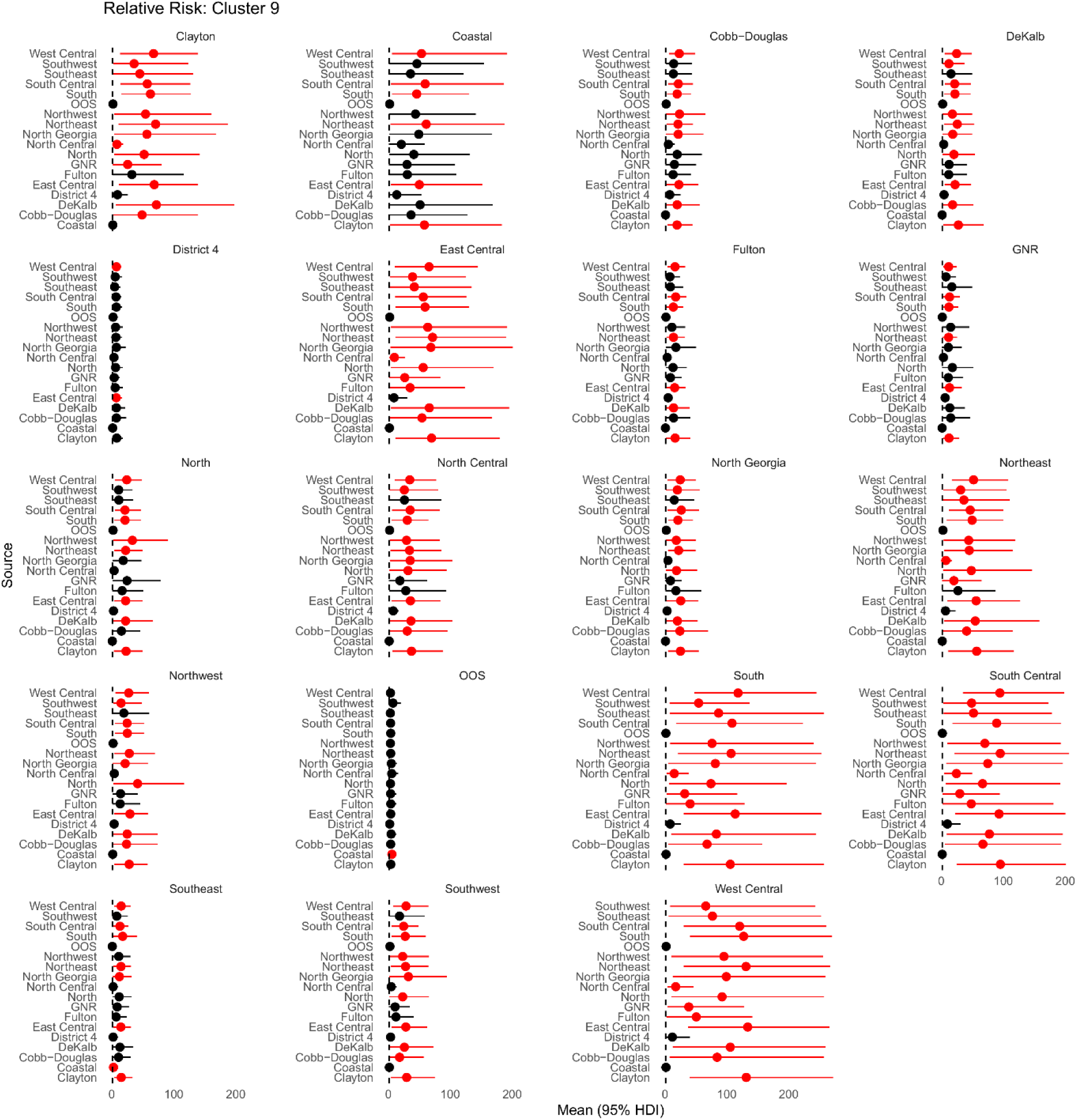
Relative risk as a viral source, Cluster 9. Forest plot depicting the relative risk of each public health district as a source for a specific sink. Relative risk is calculated as the ratio of the proportion of the times a district serves as a source for the given sink versus the proportion of times serves as a source for all other sinks. Confidence intervals excluding 1 are colored red, indicating strong evidence that the district is a significantly more likely source for the sink compared to all other possible sinks. Public health districts may have similar RR estimates for multiple sinks, indicating the district’s relatively consistent role across different sinks. Greater similarity in point estimates and overlapping confidence intervals suggest less heterogeneity among sources, indicating no significant difference in their contributions to the sink. Wide HPD intervals indicate statistical uncertainty for a particular public health district to be a source for this sink, likely due to low numbers of introductions to some locations.

**Supplementary Figure 13.**
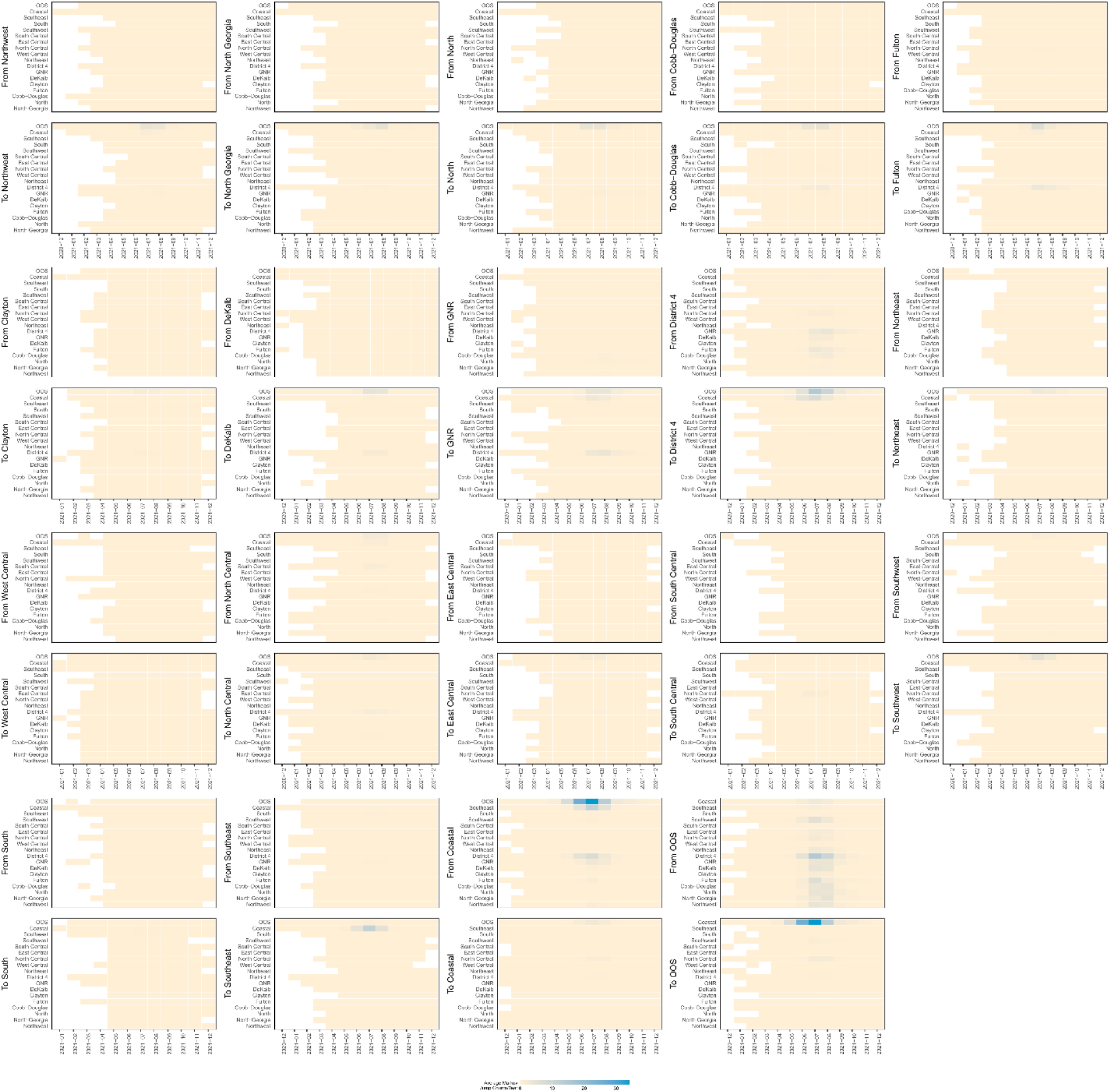
Markov jumps over time, Cluster 34. Phylogenetic discrete trait analyses inferred transitions (Markov jumps) between public health districts over time along phylogenetic branches. Darkness of each cell is proportional to the absolute number of transitions from the first column to the second column over time.

**Supplementary Figure 14.**
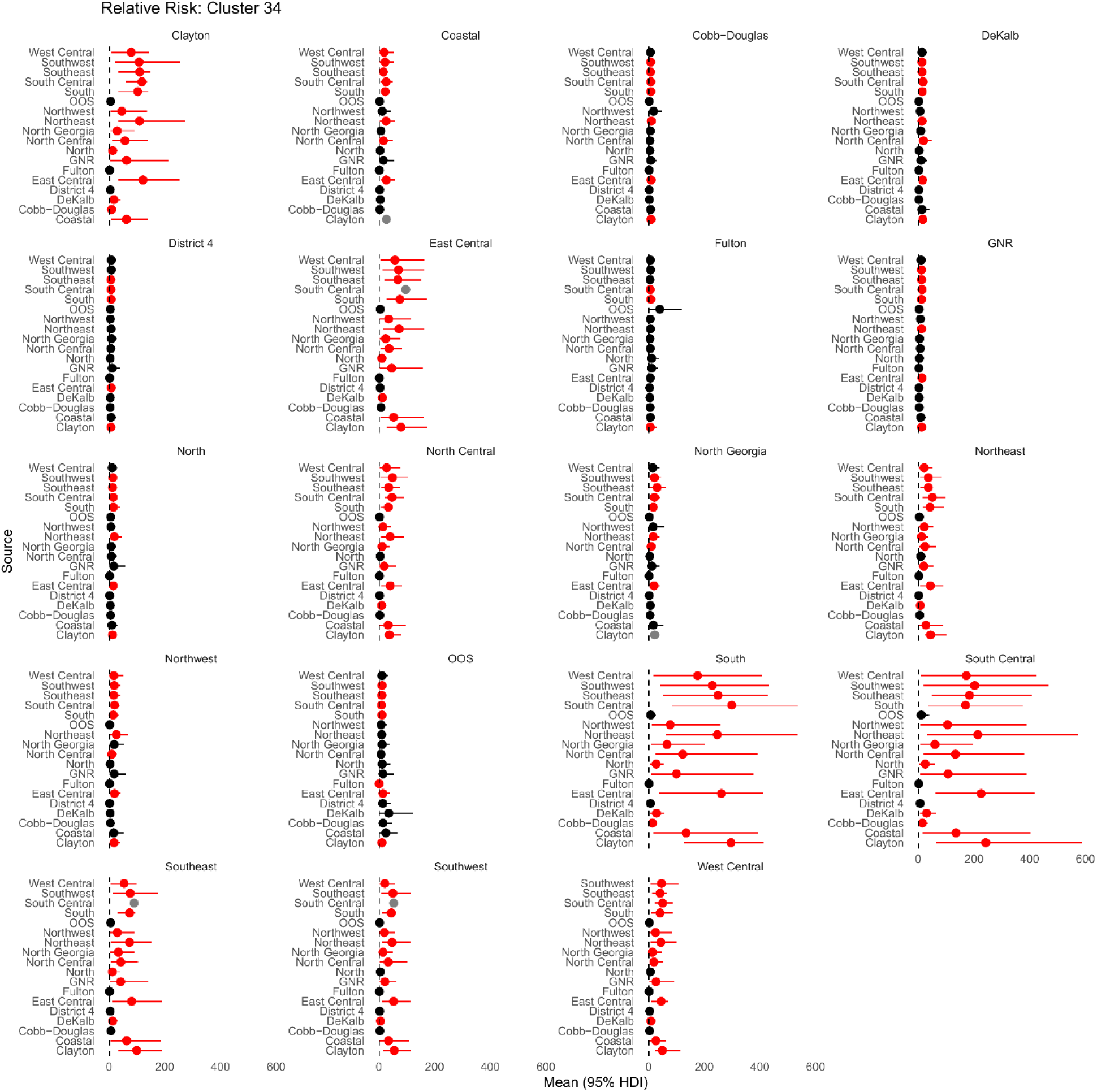
Relative risk as a viral source, Cluster 34. Mean relative risk (RR) ratios for each public health district to act as a viral source estimated from the total number of Markov jumps from or to each discrete state.

**Supplementary Figure 15.**
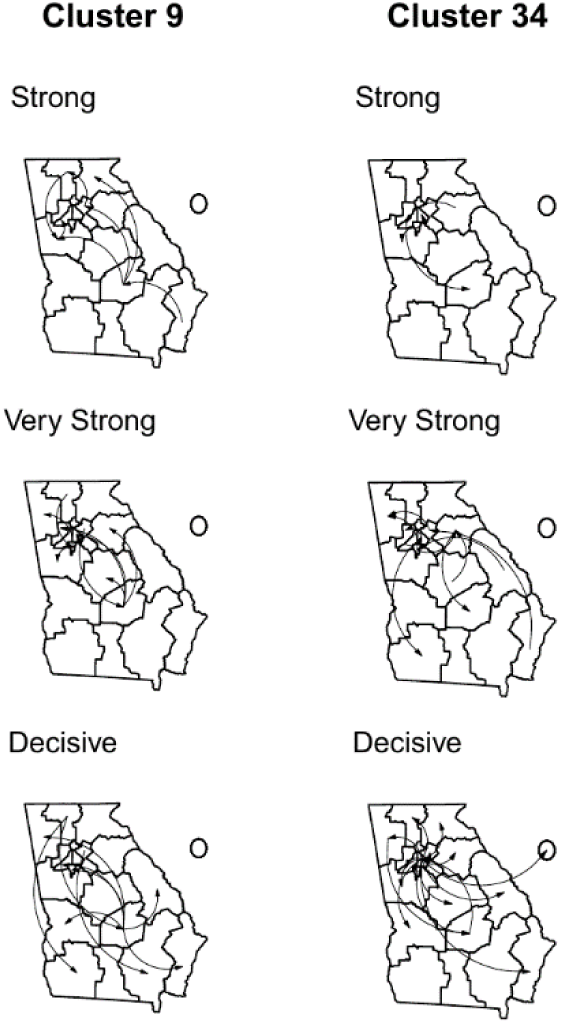
Cluster 9 and 34 statistically supported BSSVS transition rates (PP > 50%), by level of BF support (strong, very strong, decisive). Circle represents out of state.

**Supplementary Figure 16.**
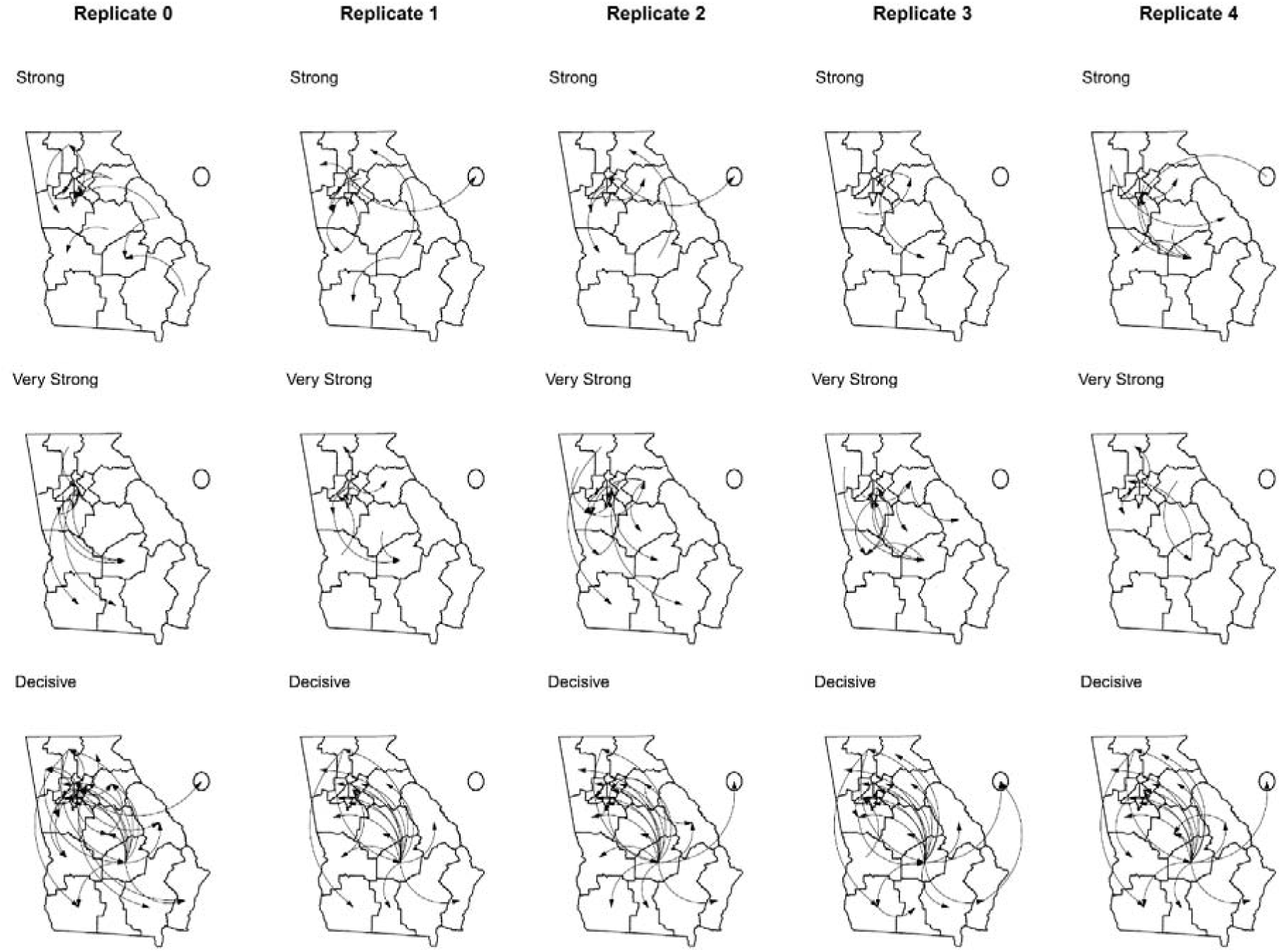
Statistically supported BSSVS transition rates (PP > 50%) by level of BF support (strong, very strong, and decisive), across subsamples. Circle represents out of state.

**Supplementary Figure 17.**
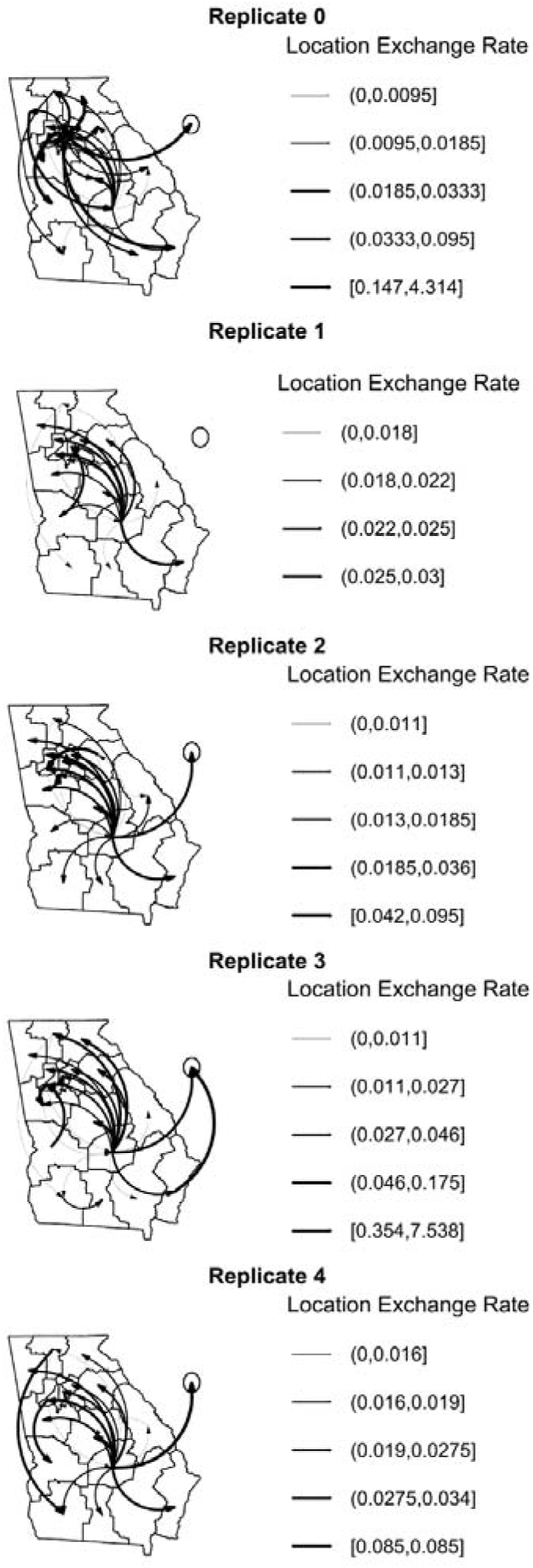
Decisive (BF > 100) and statistically supported (PP > 50%) BSSVS transition rates across subsamples, by rate quantile. Line thickness corresponds to quantile (0-25%, 25-50%, 50-75%, 75-100%, and outliers). Outliers (outside of 1.5x IQR) were removed prior to calculating transition rate quantiles and are presented separately to highlight particularly high transition rates. Circle represents out of state.

